# A Toolkit for Health-economic Analysis with Language-Agnostic Simulation and Sensitivity Analysis (THALASSA)

**DOI:** 10.64898/2026.10.17.26365802

**Authors:** David Gómez-Guillén, David Fernández-López, Jesus Cerquides, Josep Lluís Arcos, Mireia Díaz

## Abstract

**Background:** Cost-effectiveness analyses (CEA) built on simulation models are usually delivered through project-specific scripts. Each new model, and often each new question, rebuilds the same scaffolding: parameter management, calibration, base-case analysis, sensitivity analysis and export. This is costly, and is where traceability between an estimate and the model version producing it is lost. We present THALASSA, a web application providing this infrastructure once, for any model satisfying a small programming contract.

**Methods:** A simulation model is connected by one file implementing a fixed contract of R functions that determine its behaviour. THALASSA exposes the complete CEA workflow in a browser: a generated overview of the connected model, parameter editing, calibration, base-case comparison with the efficiency curve, and deterministic and probabilistic sensitivity analysis at a willingness-to-pay threshold, each with timestamped file export carrying the parameter values it was run with. Every session is bound to a repository version, so results carry their generating model version.

**Results:** We demonstrate THALASSA on two public models of contrasting difficulty: a simple tutorial model and a generic cancer screening model with two competing natural-history pathways and a 21-dimensional calibration deliberately built to be more complex. Both ran on the same unmodified setup, implementing the same contract, with no code changes.

**Conclusions:** Separating analysis infrastructure from model logic makes the CEA workflow reusable across projects and reproducible by construction. A modelling team can move from a versioned repository to audit-ready, exportable CEA outputs without writing analysis code. The integration effort does not grow with model size and one installation serves unrelated projects.

## 1 Introduction

Healthcare resources are finite, and in many health systems their allocation is guided by formal economic evaluation. Cost-effectiveness analysis (CEA) compares alternative interventions on the basis of differences in costs and health outcomes. The incremental cost-effectiveness ratio (ICER) is commonly used to express the additional cost per additional unit of health gained [1]. Vhen the relevant costs and health outcomes cannot be observed directly over the required time hori:ton, decision-analytic simulation models are used to project them, often to estimate long-term outcomes such as quality-adjusted life years (QALYs). CEA informs reimbursement decisions, the prioritisation of prevention programmes and the design of screening strategies [2].

Model-based CEA involves more than running the simulation model. Parameters must be managed. Because estimates drawn from heterogeneous literature or expert consensus rarely match the target population, the model must often be calibrated so that its outputs reproduce observed epidemiological quantities such as age-specific cancer incidence [3]. Alternative strategies must be compared, and guidelines require that uncertainty be assessed through deterministic and probabilistic sensitivity analyses [4]. The results must then be reported, and they must remain traceable to the model and inputs that generated them.

Although these methodological components are well established, the infrastructure used to implement them in day-to-day practice is less standardised. Analyses are often managed through project-specific scripts that are closely tied to one model and are difficult to reuse across projects. This creates three practical limitations for modelling teams. First, the analysis infrastructure is rebuilt for each new model or research question, which duplicates effort. Second, reproducibility depends on a clear link between results, model versions, parameter values and analysis settings [7, 8]. With scripts, that link rests on the analyst s discipline rather than on the tooling. Third, script-based workflows require programming expertise. Clinicians, health economists and decision makers therefore cannot explore alternative assumptions or scenarios themselves, and each request waits for the few team members able to run the code. These limitations point to the need for reusable analysis infrastructure that can serve different simulation models, preserve the provenance of results and make the analyses available through an accessible interface.

We present THALASSA, an R Shiny [9] application that supplies this infrastructure once, for any simulation model that satisfies a small programming contract, and exposes the complete CEA workflow in a browser. This article describes that workflow from the perspective of the end user and demonstrates it on three models: a minimal model that serves as a tutorial, a larger model with a harder calibration problem, and a real screening model used in a short research example. The aim is to show that the same unmodified application can support the complete CEA workflow across different models without model-specific analysis code.

## 2 Methods

### 2.1 THALASSA architecture and design

THALASSA separates two things that are usually entangled: the *mode*, which encodes disease natural history, strategies, costs and effects, and the *analysis infrastructure*, which differs little from one model to the next. The model lives in its own Git repository and is not modified by the infrastructure. The infrastructure supplies everything else and is not specialised to any one model.

The design principle governing that split is to ask the model author for as little as possible about how the model works. THALASSA needs to know which parameters exist, which strategies are compared and what a run returns, and it deliberately asks nothing about disease structure, cycle length, internal state or implementation language. The intent is that connecting a model should be a small, bounded task for the modelling team rather than a project of its own.

### 2.2 Model integration contract

Integration consists of adding one file, `thalassa_interface.R`, to the model repository. That file must only implement the functions specified in Table 1. The contract fixes only what the application asks for and what it receives back, leaving everything on the model side to the discretion of the model author. Model agnosticism therefore entails language agnosticism. The interface file needs only some means of invoking the model, whether directly in R or through a bridge such as `reticulate`or `Rcpp`. The effort of connecting a model is front-loaded onto the few required functions and other optional functionality, such as calibration, can be added later if needed.

**Table 1:**
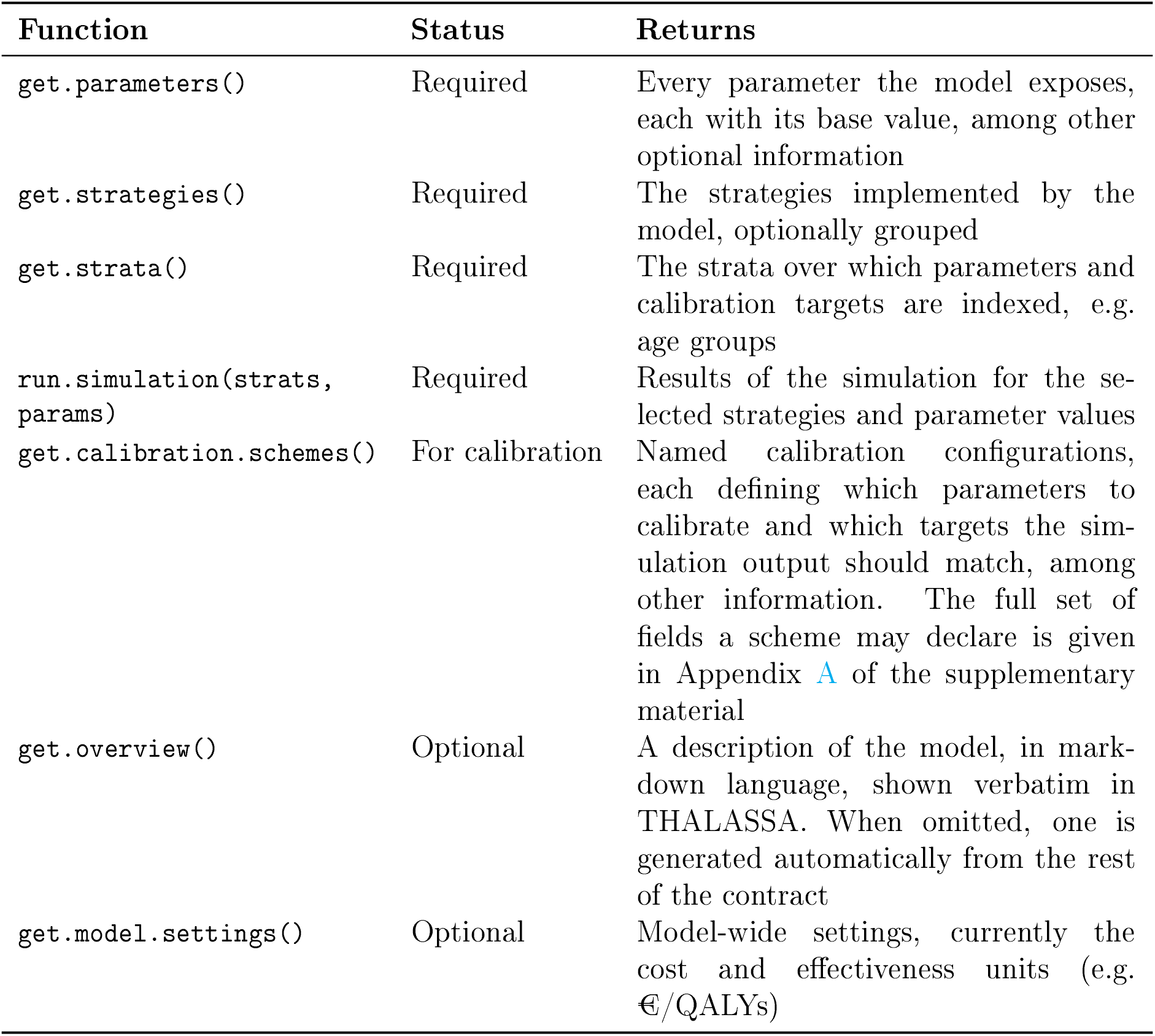
The model integration contract, as currently defined.

| Function | Status | Returns |
| --- | --- | --- |
| `get.parameters()` | Required | Every parameter the model exposes, each with its base value, among other optional information |
| `get.strategies()` | Required | The strategies implemented by the model, optionally grouped |
| `get.strata()` | Required | The strata over which parameters and calibration targets are indexed, e.g. age groups |
| `run.simulation(strats, params)` | Required | Results of the simulation for the selected strategies and parameter values |
| `get.calibration.schemes()` | For calibration | Named calibration configurations, each defining which parameters to calibrate and which targets the simulation output should match, among other information. The full set of fields a scheme may declare is given in Appendix A of the supplementary material |
| `get.overview()` | Optional | A description of the model, in mark-down language, shown verbatim in THALASSA. When omitted, one is generated automatically from the rest of the contract |
| `get.model.settings()` | Optional | Model-wide settings, currently the cost and effectiveness units (e.g. €/QALYs) |

A running demo of THALASSA, connected to the two case-study models described below, is available and linked from the project documentation at https://david-gomez-guillen.github.io/thalassa.

### 2.3 The user workflow

Once a model is connected, THALASSA presents the analysis as six sequential steps, each a tab in the browser. In each step the user makes a few selections and runs the analysis with one button, without writing code. The order reflects how a CEA is actually assembled, and most steps build on the state left by earlier ones. Figure 2 shows how the analysis steps follow one another. The steps are described below in three groups.

**Figure 1:**
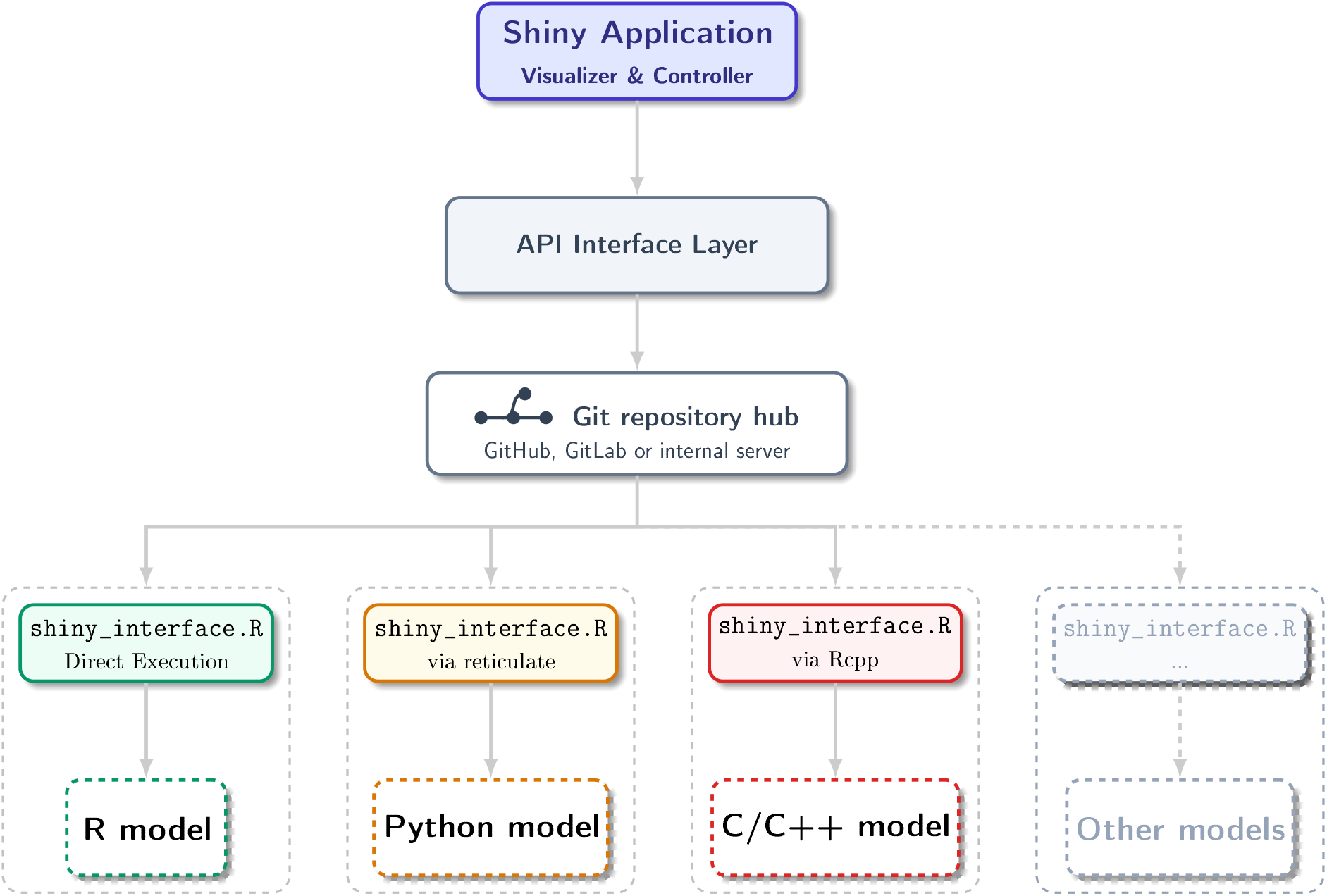
THALASSA architecture. THALASSA communicates with any simulation model exclusively through the `thalassa_interface.R`contract. Each dashed container is one model repository, holding the model and its interface file together, and is loaded at run time at a user-selected branch or tag. The contract constrains only the signatures of the functions it declares, not the model behind them, so the model may be implemented in any language the interface file is able to invoke.

**Figure 2:**
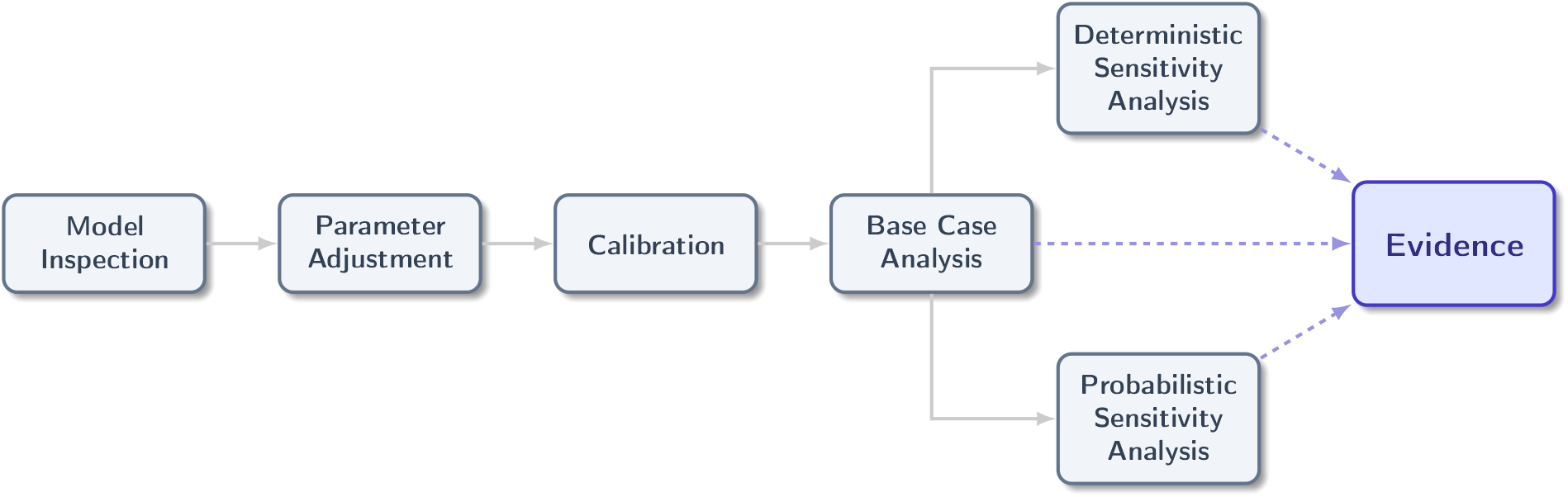
The analysis workflow of THALASSA, once the model and its version have been selected. Model inspection, parameter adjustment and calibration precede the base case analysis, and each step is carried into the next. The deterministic and the probabilistic sensitivity analysis both start from the base case and run independently of each other. The base case and the two sensitivity analyses are what produce the evidence the session is run for.

#### 2.3.1 Model selection and inspection

At session start the user chooses a model and then the version of it to run. The version selector lists the versions published in the model repository. These are either named releases, such as a version submitted with a paper, or lines of ongoing development. This single choice fixes the provenance of everything produced in the session.

The Overview tab then shows the description the model declares for itself, written by the model author, which is where the structure, the data sources and the meaning of the parameters belong. A model that declares none is described anyway: THALASSA generates the overview from the rest of the contract, describing the strategies, parameters, strata and every calibration scheme. This is the step at which a user can get acquainted with the model. Figure 3 shows the tab for the tutorial model of case study 1.

**Figure 3:**
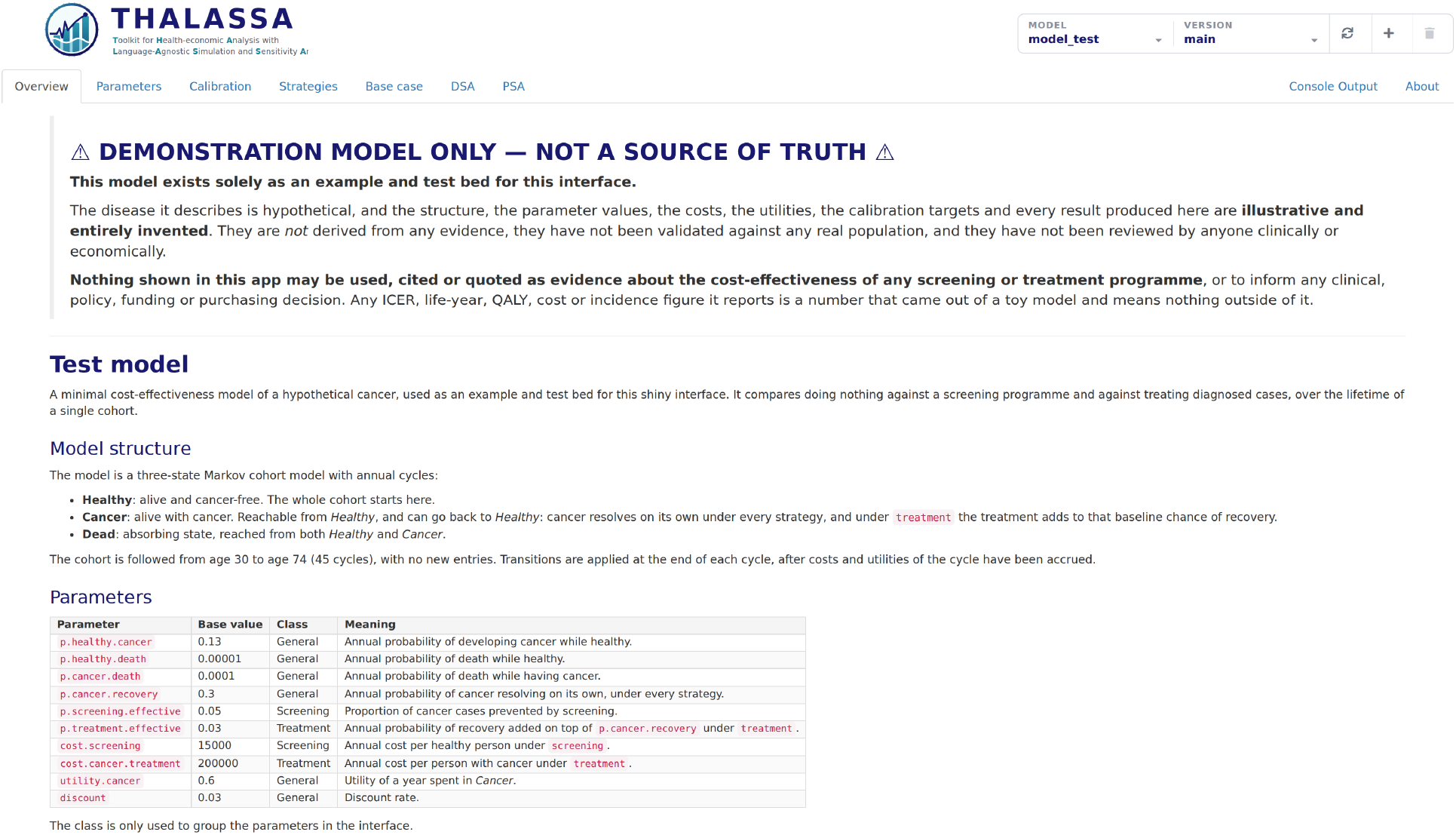
The Overview tab, here for the tutorial model of case study 1. Because that model implements `get.overview()`, the tab renders the description its author wrote.

**Figure 4:**
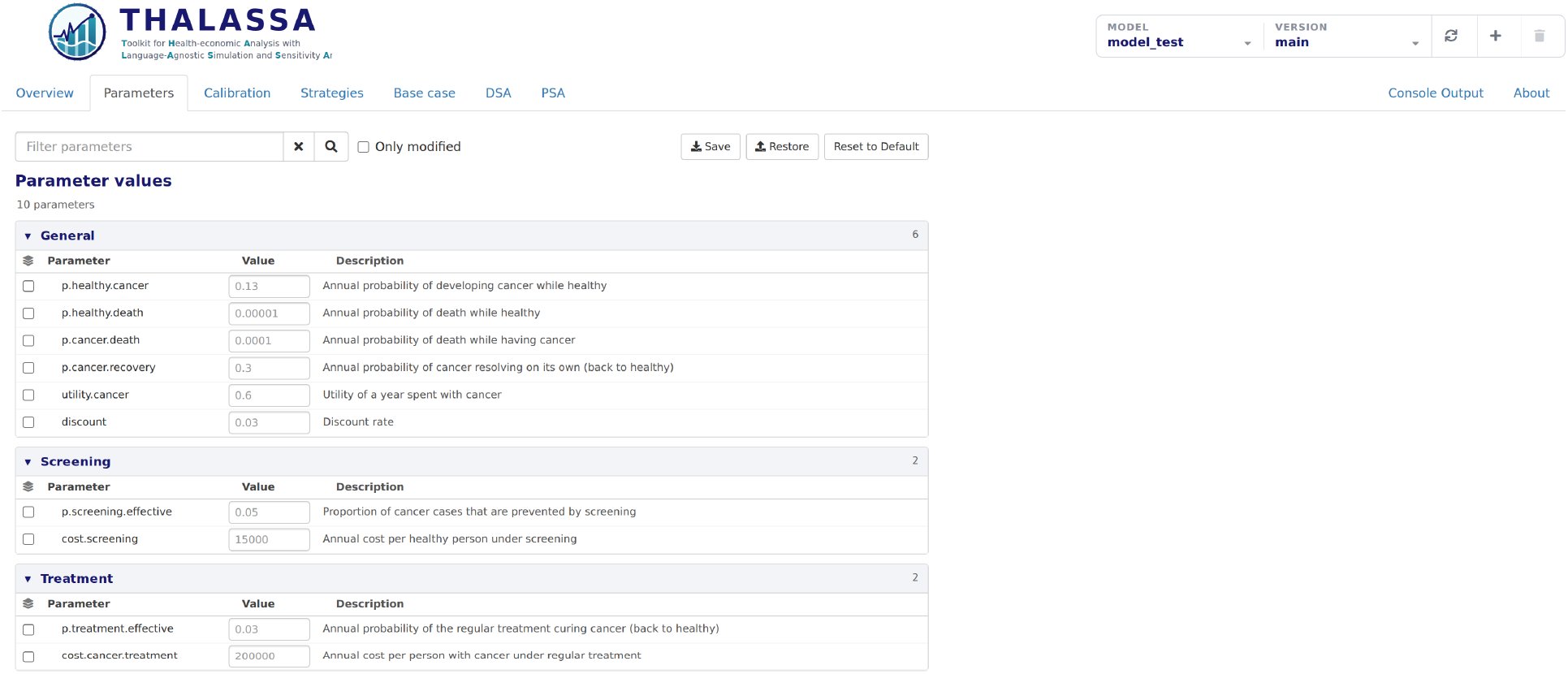
The Parameters tab, for the same model. The grid is grouped into the classes the model declares, here general, screening and treatment, each collapsible and reporting how many parameters it holds. The save and restore buttons write and read the parameter set as a workbook.

#### 2.3.2 Parameter management and calibration

The Parameters tab presents every parameter the model exposes as a grid of collapsible sections, one per class, such as costs, probabilities and utilities, with a stratified parameter folding away its strata under its own row. These parameters can be searched, filtered and reset to default values. The values set in this tab propagate to all downstream steps of the CEA workflow like the base-case analysis, DSA or PSA.

The Calibration tab lists the schemes declared by the model that the user can choose. A scheme is a named calibration predefined group of settings, stated by the model author, that fixes which parameters are being calibrated, which observed targets the simulation output is fitted to and in which strata, where the search starts and how the discrepancy between model and target is measured. A model may declare several schemes to satisfy different calibration needs. A scheme may also declare a feasible region, in which case the tab states in text what that region requires, can restrict the search to it, and marks on the error plot the simulations that fell outside it. Progress is displayed live: current error, best-so-far trace, and modelled versus observed curves, with the elapsed time and the best error reported when the run ends. The resulting parameter set can be fixed for the remaining CEA steps, so that the base case and both sensitivity analyses run on calibrated values without further configuration.

The calibration engine provides classical optimisation methods, such as Nelder-Mead and simulated annealing, but favours Bayesian optimisation (BO), whose greater sample efficiency is advantageous when individual simulations are computationally expensive. Any optimisation method can be combined with a learned low-dimensional latent space that keeps proposals within clinically plausible regions [10, 12]. Finally, an agent-driven proposal method is offered, in which a large language model proposes each next parameter vector. Methodological development of the calibration engine itself is reported separately [10, 11]. Appendix B of the supplementary material describes the available methods in more detail.

#### 2.3.3 Base-case and sensitivity analyses

The Base Case tab compares selected strategies, chosen through a hierarchical tree for large strategy sets, and reports total costs and effects per strategy together with the cost-effectiveness efficiency frontier. Alongside the ICER the table reports the net health benefit (NHB) at a user-defined willingness-to-pay threshold.

The deterministic sensitivity analysis (DSA) tab varies each selected parameter around its value by a user-specified percentage and produces a tornado chart ranked by impact on either the ICER or the NHB at the chosen threshold. The probabilistic sensitivity analysis (PSA) tab draws from the parameter distributions declared by the model and produces the cost-effectiveness plane scatter, with the threshold drawn on it, and the acceptability curves. Both run against calibrated values if calibration is active. Plots are interactive, each point reporting its strategy, costs and effects on hover.

#### 2.3.4 Reproducibility and result provenance

Throughout all six steps, every analysis tab exports its results as a timestamped spreadsheet carrying a sheet with the full parameter set the run used, flagging which of those values came from a calibration, and a further sheet recording the model, the version and the settings the run was set up with. That same file is read back by the tab that wrote it, which puts the results and those settings on screen again without calling the model, and the Parameters tab writes and reads the parameter set in the same way. A console tab streams the model’s own output, so long runs remain inspectable and model-side errors are visible to the user. Appendix E of the supplementary material details the exported workbooks and how a result is reproduced from them.

As a result, several properties that are normally the analyst s responsibility become structural. Table 2 maps these properties to items of established modelling good-practice and reporting guidance.

**Table 2:**
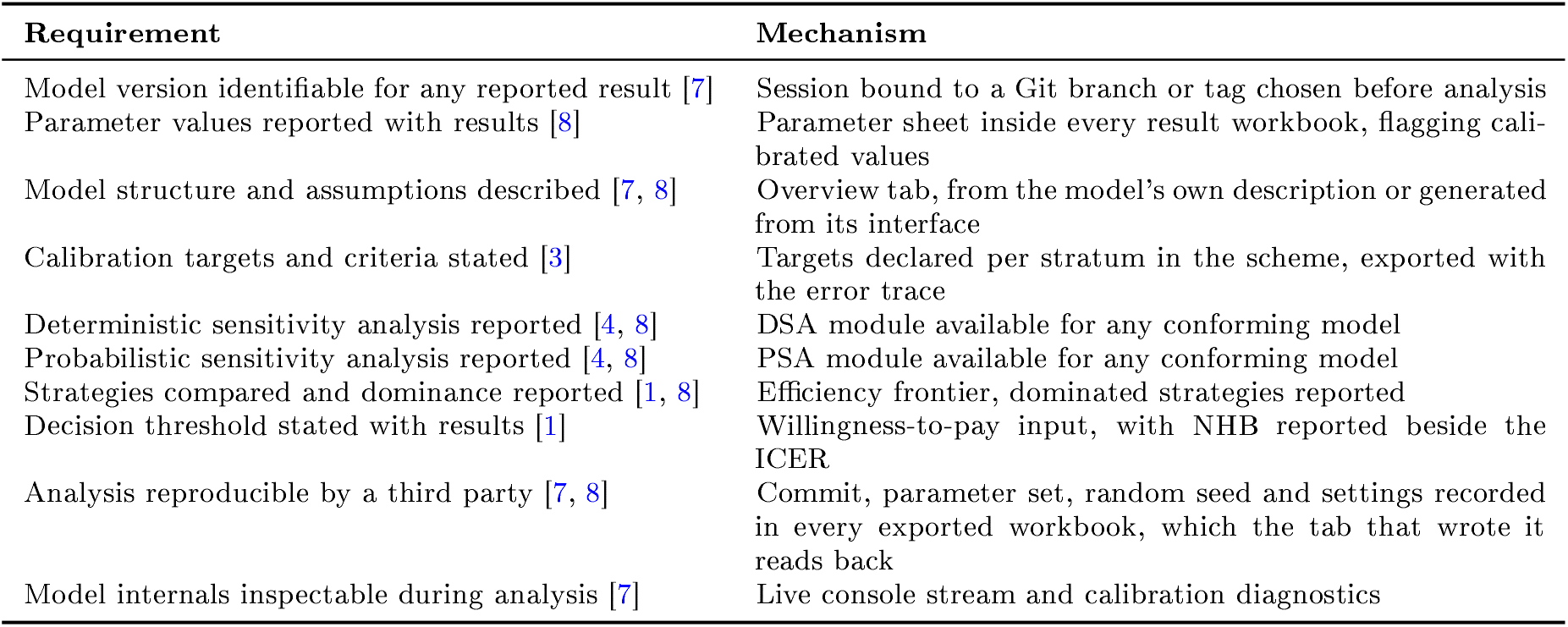
Selected modelling good-practice and reporting requirements, and how THALASSA supports them. The full mapping, including the requirements that remain the analyst’s responsibility, is in Appendix F of the supplementary material.

### 2.4 Demonstration models and case-study design

THALASSA was demonstrated on two publicly available simulation models of contrasting complexity, summarised in Table 3. The first is a minimal tutorial model designed to illustrate the complete workflow. The second is a more complex synthetic cancer screening model with two competing natural-history pathways and a higher-dimensional calibration problem, designed to test the application under greater analytical complexity. It was written to be hard to calibrate rather than to mimic clinical reality.

**Table 3:** The two case-study models. Both were driven from the same unmodified installation of THALASSA, both satisfy the same contract of Table 1, and both are publicly available.

|  | Case study 1 | Case study 2 |
| --- | --- | --- |
| Question | Synthetic screening and treatment example | Synthetic cancer screening with two competing natural-history pathways |
| Model type | Markov cohort, 3 states, annual cycles, age-stratified | Markov cohort, 13 states over two parallel lesion routes, annual cycles, age-stratified |
| Repository | `cea-model-test`<br>https://github.com/david-gomez-guillen/cea-model-test | `cea-model-test2`<br>https://github.com/david-gomez-guillen/cea-model-test2 |
| Age strata | 9 five-year groups, ages 30 to 74 | 7 ten-year groups, ages 20 to 89 |
| Strategies | 3, ungrouped | 4, grouped under three screening modalities: no screening, a biennial screening test, and a screening procedure every ten years from 50 or from 45 |
| Parameter entries | 10 | 64 |
| Parameter classes | 3, declared as general, screening and treatment | 9, separating the two natural-history routes from cancer progression, survival, each screening modality, costs and utilities |
| Calibration target | Age-specific cancer incidence, 9 strata | Three series at once, cancer incidence, lesion prevalence and late-stage share at diagnosis, over 6 strata |
| Free calibration parameters | 9, annual healthy-to-cancer probability by age stratum | 21, the onset probability of each lesion type and the progression probability of high-grade lesions, each by age stratum |
| Calibration schemes declared | 1 | 2, one unconstrained and one adding clinical plausibility constraints |
| Calibration difficulty | One free parameter per stratum, coupled across strata | Coupled across strata and routes, several local minima |

Neither model has clinical validity, and no result below is a statement about any cancer or about any screening programme. The base values are synthetic, but the implementation is not. Both models are complete Markov cohort simulations with age stratification, competing transitions, per-cycle cost and utility accumulation and discounting, written as a model for a real evaluation would be. THALASSA drives them through the same contract and the same code paths it would use for such a model. Both models are public, at https://github.com/david-gomez-guillen/cea-model-test and https://github.com/david-gomez-guillen/cea-model-test2, so a reader can verify and replicate every analysis.

The tutorial model, kept in the repository `cea-model-test`, is a three-state Markov cohort model, healthy, cancer and dead, run in annual cycles from age 30 to 74 and reported over nine five-year strata. It compares three strategies, `no_intervention`, `screening`and `treatment`. It declares one calibration scheme, which adjusts the annual healthy-to-cancer probability in each of the nine strata to fit a synthetic incidence curve. This case study runs the complete workflow in a single session, from model selection to probabilistic sensitivity analysis. Two calibration methods were compared, Nelder-Mead and Bayesian optimisation. The base case and both sensitivity analyses used a willingness-to-pay threshold of 25,000 «/QALY. The sensitivity analyses compared screening against no intervention over the screening parameters, each varied by *±*50%, and the probabilistic analysis used 1,000 samples.

The second model, kept in the repository `cea-model-test2`, is a synthetic cancer screening model in which cancer arises through two parallel routes. A slow pathway carries most cases through a low-grade and then a high-grade lesion, and a fast pathway carries a minority of them quickly, through a single lesion type. Each route has its own pair of undiagnosed cancer states, because fast-pathway cancers reach an advanced stage sooner and stay asymptomatic longer. The model compares no screening against a biennial screening test from 50 to 74 and against a screening procedure every ten years starting at 50 or at 45. The calibration difficulty arises from the interaction between the two routes, whose separate contributions to the observed targets are not easily disentangled.

The model declares two calibration schemes over the same 21 values. The unconstrained scheme, shown in the tab as *Lesion onset and progression*, calibrates the onset probability of each lesion type and the progression probability of high-grade lesions across the seven strata. It fits them to three target series at once, cancer incidence, lesion prevalence and late-stage share at diagnosis. Its solutions need not be clinically plausible. The constrained scheme, *Constrained onset and progression*, adds two requirements. Each of the three probabilities must be non-decreasing in age, and total fast-pathway lesion onset must stay below 35% of total low-grade lesion onset. The scheme states these requirements as a function returning the violation of each one, and also as plain text, which the tab shows beside the scheme and the agent-driven calibrator reads.

The second case study focuses on the more advanced calibration methods THALASSA offers. Four methods were compared under each scheme, all run from the same tab by a user who does not need to understand their technical details. The remaining steps of the workflow are carried out exactly as in the first case study and are not repeated. The calibration protocols of both case studies are given in Appendices C and D of the supplementary material.

A third, brief case study shows THALASSA in a real research setting. It uses `cea-anus`, an anal cancer screening model for HIV-positive men who have sex with men. The model is a Markov cohort model with 22 states and six-month cycles, run from age 40 to 80, and it offers 46 screening and treatment strategies. The research question was whether high-grade squamous intraepithelial lesions (HSIL) found after screening with a high-risk HPV E6/E7 mRNA test should be treated with infrared coagulation (IRC) or with trichloroacetic acid (TCA). The sensitivity analyses covered the five parameters specific to the two treatments, their costs and success probabilities. The DSA varied each of them by *±*20%, and the PSA drew 1,000 samples with a relative standard deviation of 10%.

## 3 Results

### 3.1 Tutorial case study: end-to-end CEA workflow

The `cea-model-test`model was selected and pinned at branch main with the two selectors at the top right of Figure 3. Those selectors remain visible throughout the session, so every screenshot in this case study states the model and version that produced it. The model implements its own overview, which describes the three states and the transitions between them, the cohort followed from age 30 to 74, the ten parameters with their base values and meanings, and what the calibration scheme fits (Figure 3). Base values were left at model defaults.

#### 3.1.1 Calibration

The scheme s declared starting point fits the synthetic incidence curve poorly (Figure 5), so calibration is needed. A Nelder-Mead search produced a suboptimal fit, and repeating the run with Bayesian optimisation fitted the target curve closely. The two runs are compared in Figure 6.

**Figure 5:**
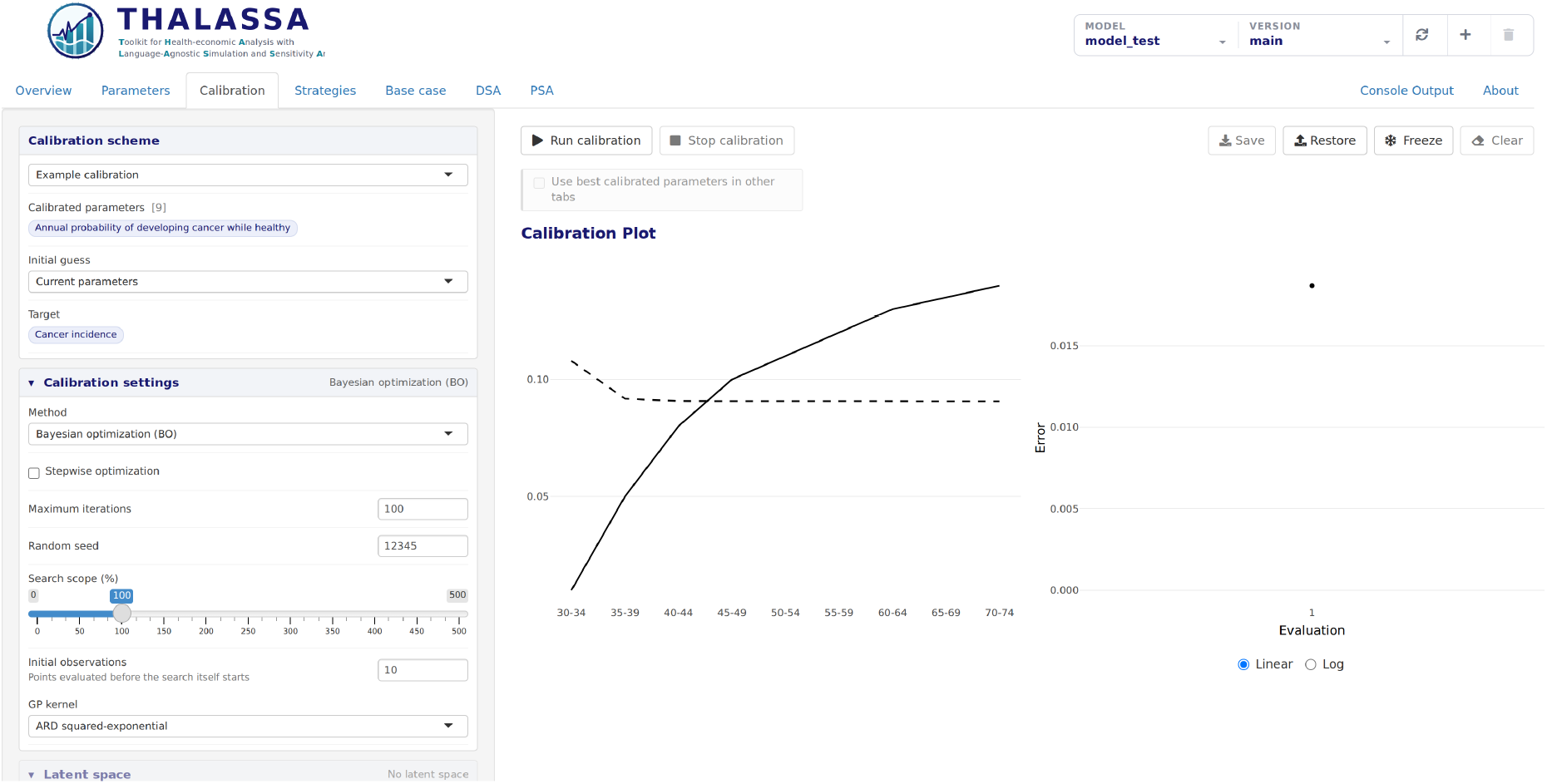
The Calibration tab of case study 1 at the scheme s declared starting point. The simulated curve shows a very poor fit to the target.

**Figure 6:**
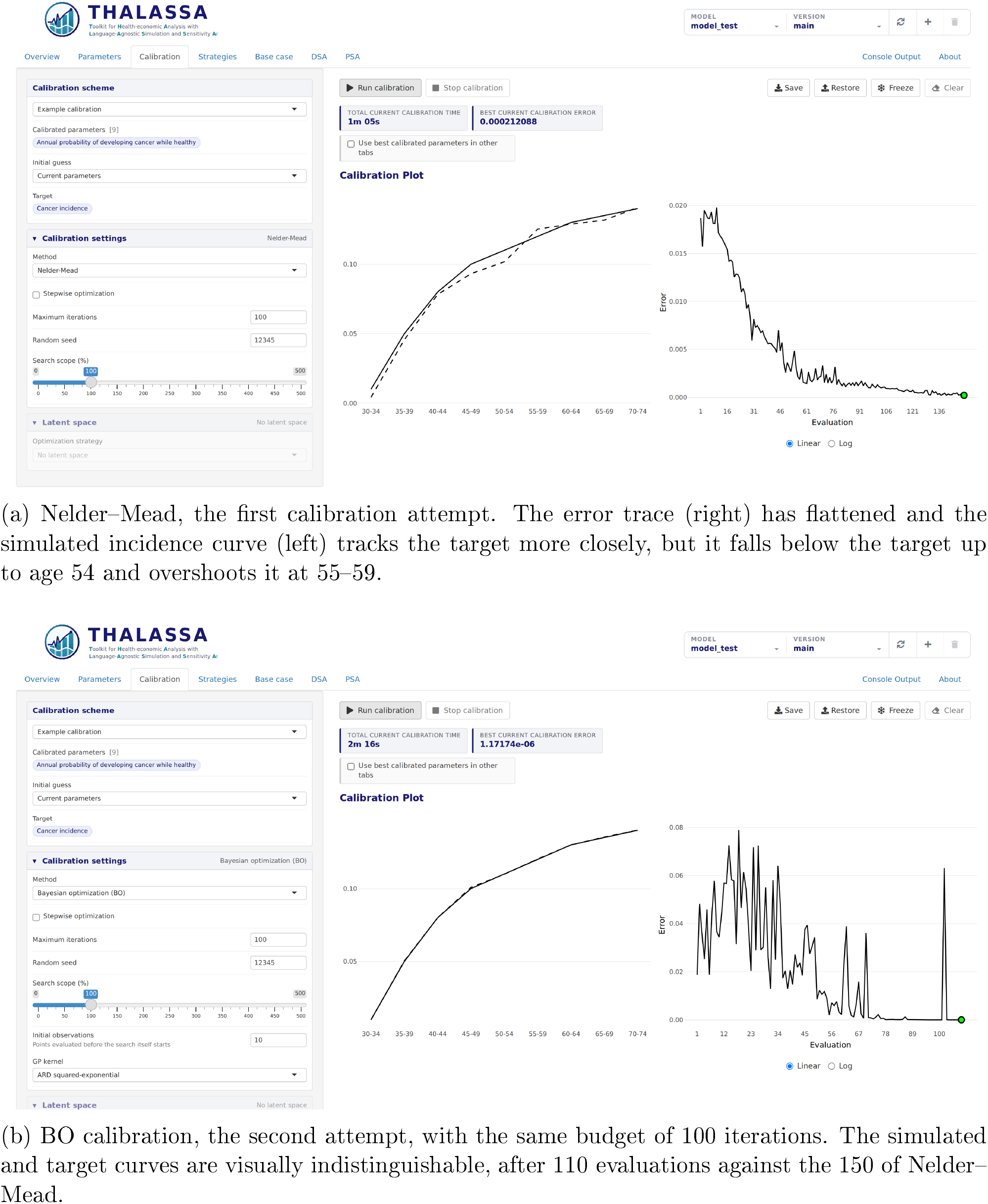
The Calibration tab of case study 1 at the end of each run. Each panel shows the scheme and the method settings that produced it on the left, the elapsed time and best error above the plots, the simulated/target curves on the left and the error trace on the right.

The calibrated values from the Bayesian run were saved and *Use best calibrated parameters in other tabs* was enabled, so the remaining steps ran on those values and the Parameters tab showed them in place of the defaults.

#### 3.1.2 Base-case and sensitivity analyses

On calibrated values, the base case ran with one button press in the Base Case tab, and the three strategies produced the results in Figure 7. Screening and treatment both stand on the efficiency frontier, at ICERs of 79,560 and 408,577 € /QALY respectively. At the threshold of 25,000 € /QALY neither is cost-effective, and no intervention is the preferred strategy.

**Figure 7:**
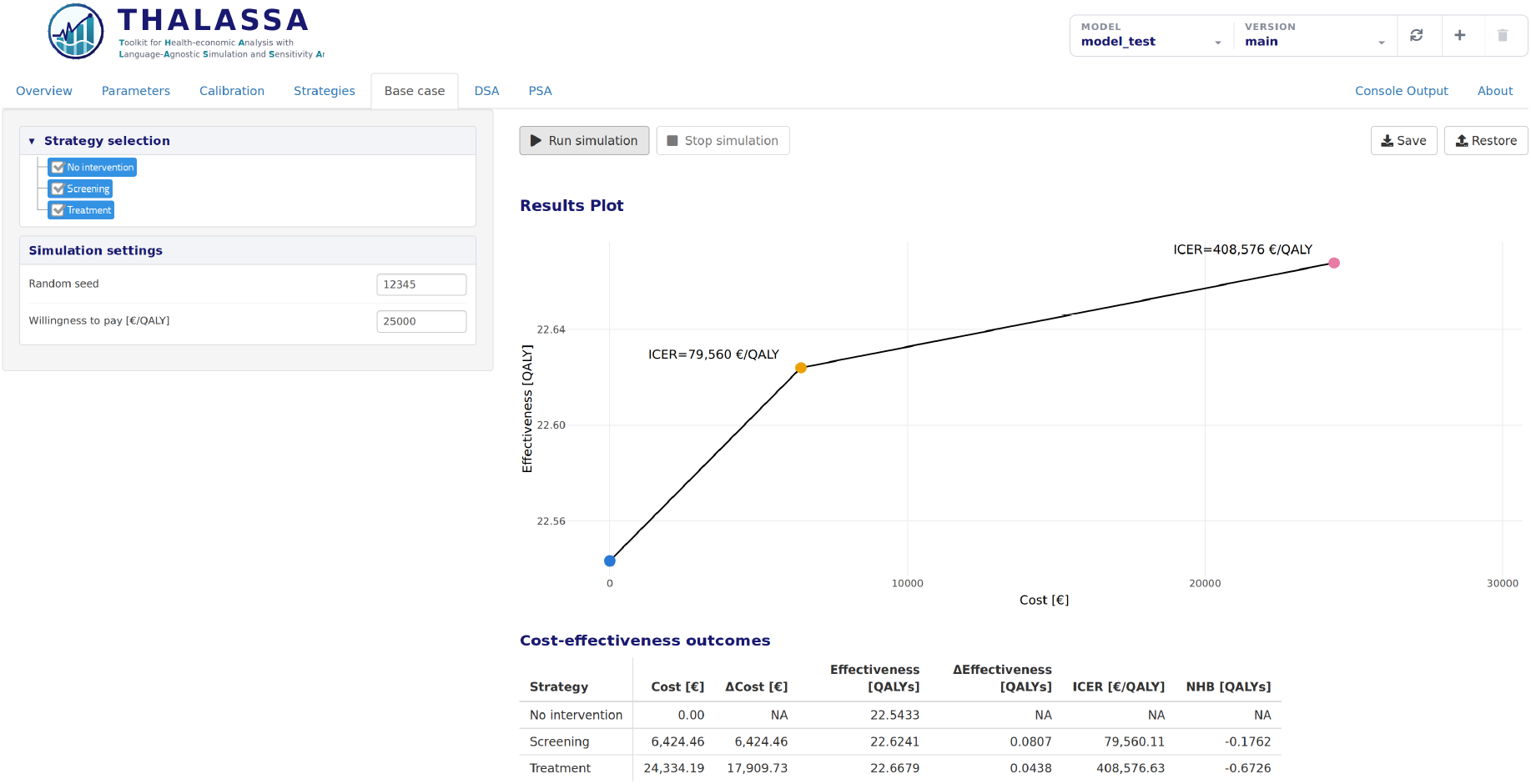
The Base Case analysis tab of case study 1 on calibrated values, all three strategies selected, as generated by THALASSA without additional post-processing. The efficiency frontier shows the no intervention, screening and treatment. The results table below the plot reports costs, effects, the ICER and the NHB at the willingness-to-pay threshold set in the sidebar.

The sensitivity analyses tested that conclusion against the uncertainty in the parameters, on the same calibrated values. The deterministic analysis (Figure 8a) showed that no single parameter brings screening below the threshold. The lowest ICER, 39,780 € /QALY, came from a screening cost of 7,500 per year, at the low end of its range. The probabilistic analysis (Figure 8b) varies the same parameters jointly. Only 9.9% of the 1,000 draws are cost-effective at the threshold, which requires both parameters to move favourably together. Both analyses therefore support the base-case conclusion that screening is not cost-effective at this threshold.

**Figure 8:**
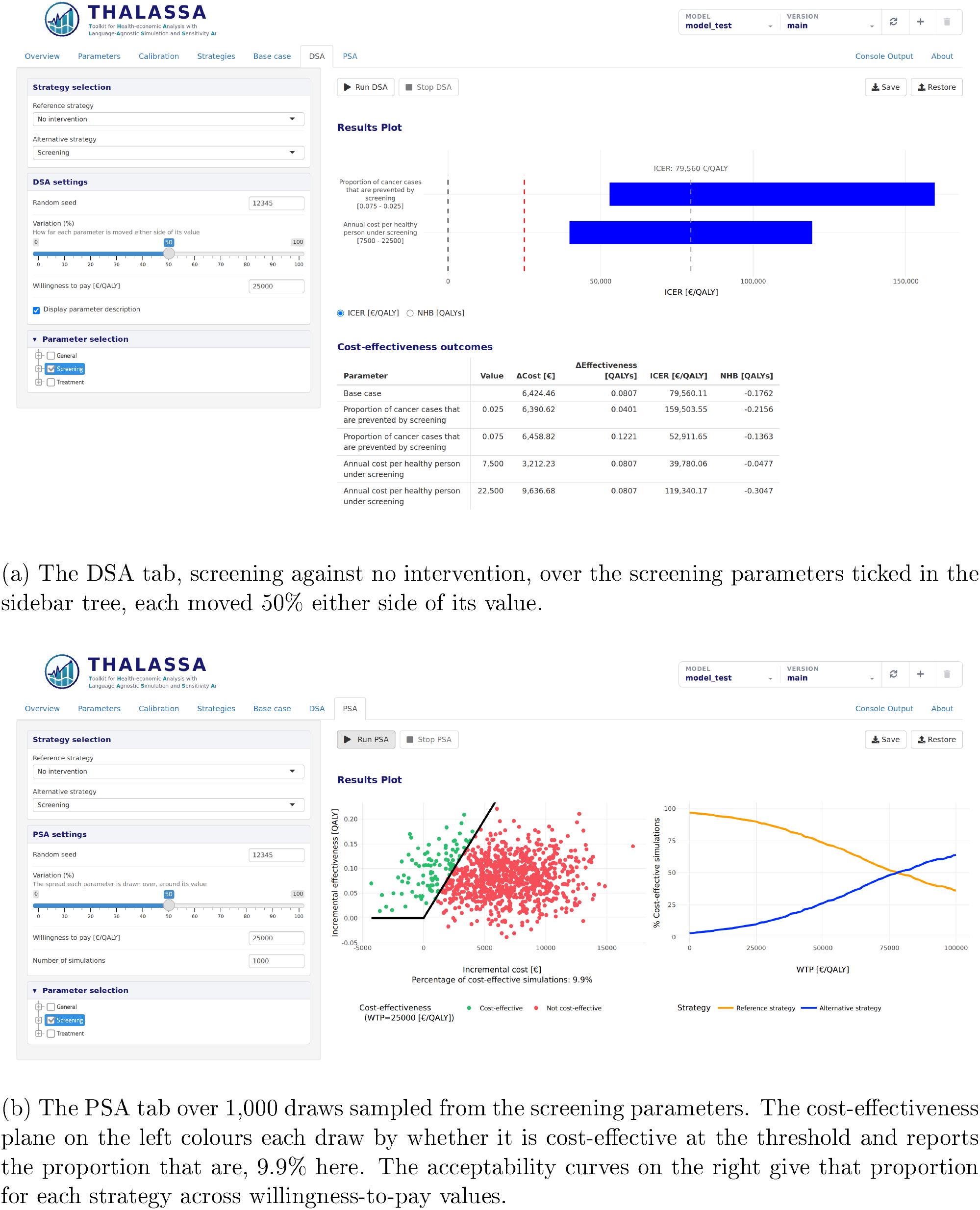
Case study 1, sensitivity analyses, as produced by the application without additional post-processing.

#### 3.1.3 End-to-end analysis and result traceability

All six steps of this case study were carried out in a single browser session, and each required only a few selections followed by one button press. Each analysis tab displayed the settings that produced its results and exported them to a workbook it can read back, so every number in Figures 6, 7, 8a and 8b is traceable to the run behind it. Those figures are everything the case study produced, shown exactly as the application generated them, with no code written and no post-processing done at any point.

### 3.2 Complex case study: advanced calibration in a higher-complexity model

This case study reports calibration for the `cea-model-test2`model, under the two schemes described in Section 2.4.

#### 3.2.1 Unconstrained calibration

Table 4 shows the four runs under the unconstrained scheme, and Figure D1 of the supplementary material shows the Calibration tab at the end of each. Nelder-Mead calibrated well but needed far more model simulations than the budget we would like to spend. Bayesian optimisation was more economical but did not calibrate as well. Its trust-region variant TuRBO made better use of the same budget and came close to the best error the targets allow. The agent-driven method, in which a large language model proposes each next parameter vector from the history of simulations and from the model source, was the most efficient. It reached a comparably good fit with a far smaller simulation budget than any other method.

**Table 4:** The four unconstrained calibration runs of case study 2. Some runs were made without the model’s artificial delay, so every elapsed time is stated at three seconds per simulation, to make the runs comparable (Appendix D of the supplementary material).

| Method | Iteration budget | # Simulations |  | Elapsed time | Best error |
| --- | --- | --- | --- | --- | --- |
|  |  | Initial | Search |  |  |
| Nelder–Mead | 1000 | 22 | 1228 | 83:46 | $1.7 \times 10^{-4}$ |
| Bayesian optimisation | 250 | 10 | 250 | 65:20 | $1.5 \times 10^{-3}$ |
| Bayesian optimisation [TuRBO] | 250 | 10 | 250 | 71:09 | $4.7 \times 10^{-5}$ |
| Agent-driven (LLM) | 25 | 1 | 24 | 216:18* | $9.1 \times 10^{-5}$ |

\* The agent-driven run’s elapsed time additionally reflects local infrastructure for the language model inference itself, better hardware or a commercial API endpoint could reduce it substantially.

#### 3.2.2 Constrained calibration

A low error is not sufficient on this model. The unconstrained runs return onset curves that fall and rise again with age (Figure D1). Such solutions can reproduce the targets, but an epidemiologist would not accept them as clinically plausible. The constrained scheme addresses this. Table 5 shows the four constrained runs, and Figure D2 of the supplementary material shows the Calibration tab at the end of each, with infeasible evaluations shaded.

**Table 5:** The four constrained calibration runs of case study 2, under the `constrained` scheme, which calibrates the same 21 values and requires each probability to be nondecreasing in age and total fast-pathway lesion onset to stay a minority of total low-grade lesion onset. As in Table 4, every elapsed time is stated at three seconds per simulation.

| Method | Iteration budget | # Simulations |  | Elapsed time | Best error |
| --- | --- | --- | --- | --- | --- |
|  |  | Initial | Search |  |  |
| Nelder–Mead, infeasible penalised | 1000 | 22 | 1298 | 89:45 | $6.2 \times 10^{-3}$ |
| Nelder–Mead in a VAE latent space | 1000 | 22 | 858 | 57:21 | $4.2 \times 10^{-4}$ |
| Bayesian optimisation [SCBO] | 100 | 10 | 100 | 36:01 | $1.6 \times 10^{-3}$ |
| Agent-driven (LLM), constrained | 25 | 1 | 24 | 262:56* | $3.0 \times 10^{-4}$ |

\* The agent-driven calibration run’s elapsed time additionally reflects local infrastructure for the language model inference itself, better hardware or a commercial API endpoint could reduce it substantially.

Nelder-Mead with infeasible solutions penalised wasted much of its budget evaluating them (Figure D2a). Wrapping the same method in a latent space avoided this. A variational autoencoder (VAE), trained on feasible parameter sets generated by the interface, learned a lower-dimensional representation of the feasible region, and the search in that space was both faster and free of infeasible proposals. SCBO, a constrained Bayesian method, learns the feasible region from the violations it observes. It evaluated a few infeasible points early in the run and almost none once its surrogate had learned the region. The constrained agent-driven calibration was as efficient as the unconstrained one.

### 3.3 Real-world case study: treatment of HSIL

The `master` version of the `cea-anus` model was selected and the base case, DSA and PSA were run in one session. In the base case (Figure 9), TCA cost € 58 more than IRC and gained 0.0118 QALYs, an ICER of 4,929 € /QALY.

**Figure 9:**
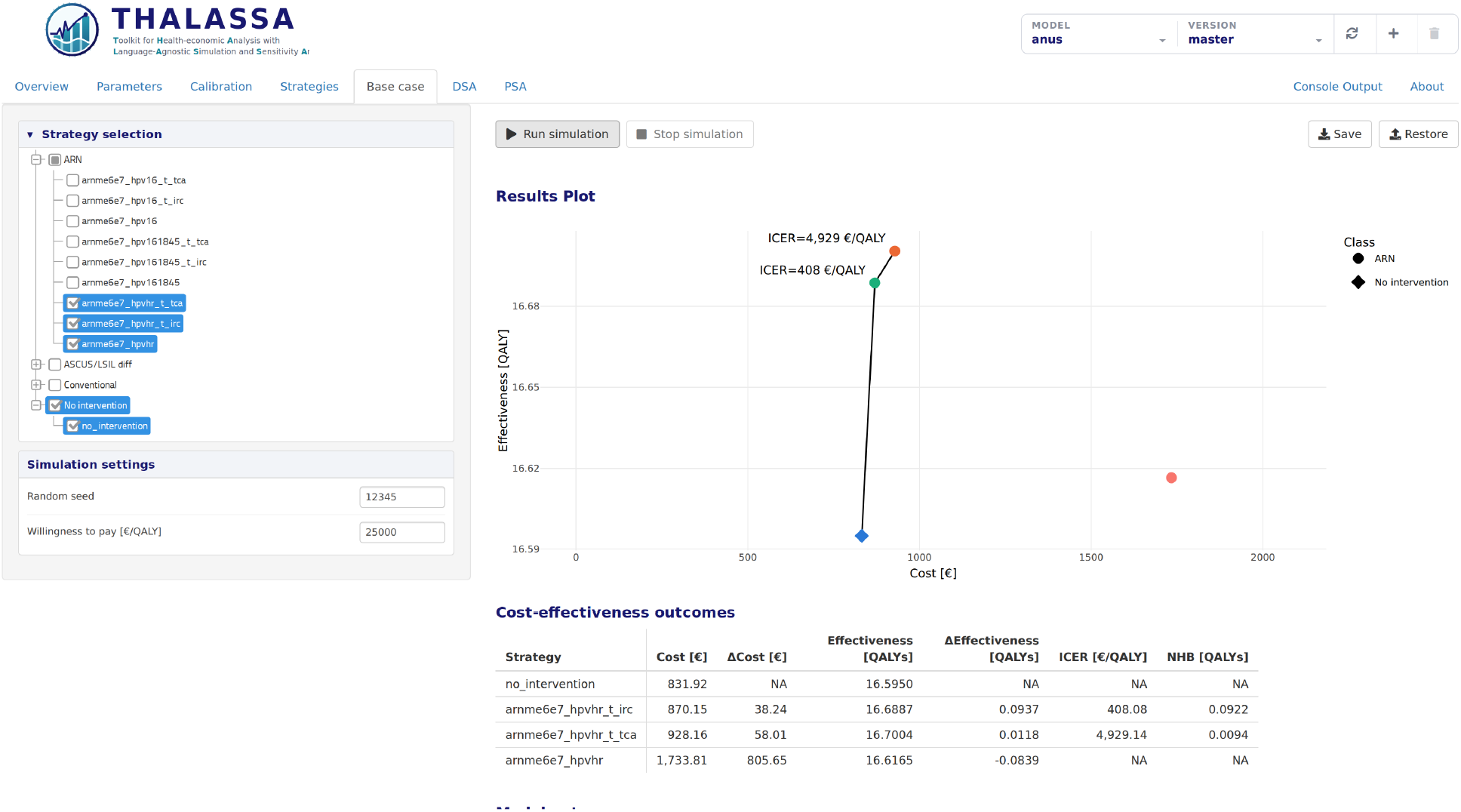
The Base Case tab of case study, with no intervention, screening with follow-up only, and screening followed by IRC or by TCA, as produced by the application without additional post-processing.

In the DSA (Figure 10), no parameter moved the ICER above the threshold. The success probability of TCA had the largest effect, with ICERs from 2,199 to 23,949 € /QALY, all below the WTP threshold.

**Figure 10:**
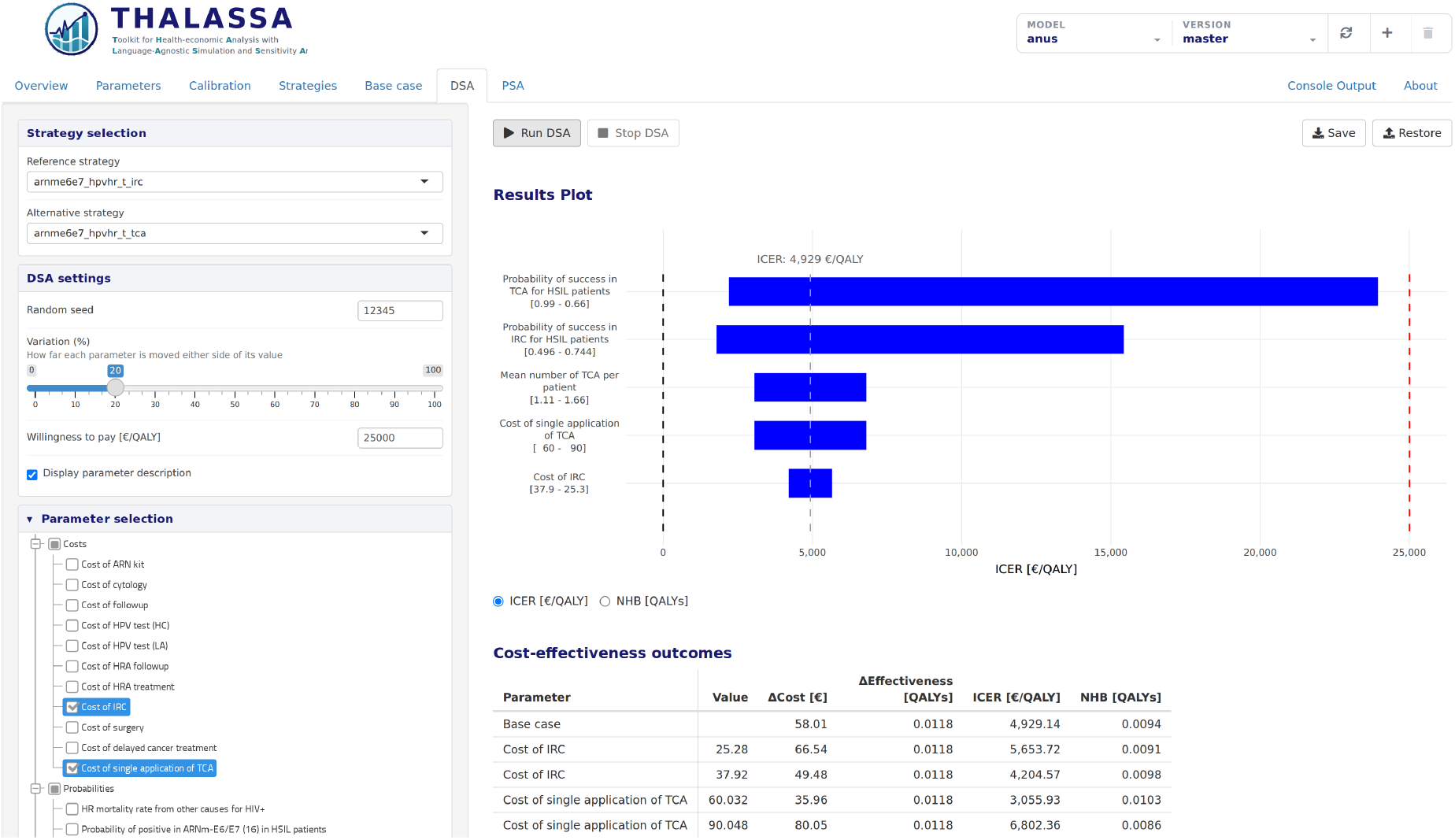
The DSA tab of case study, TCA against IRC, each treatment parameter moved 20% either side of its value.

In the PSA (Figure 11), TCA was cost-effective in 94.9% of the draws. In this population, TCA therefore appears to be the cost-effective treatment of HSIL, and the result depends mainly on how much more effective TCA is than IRC.

**Figure 11:**
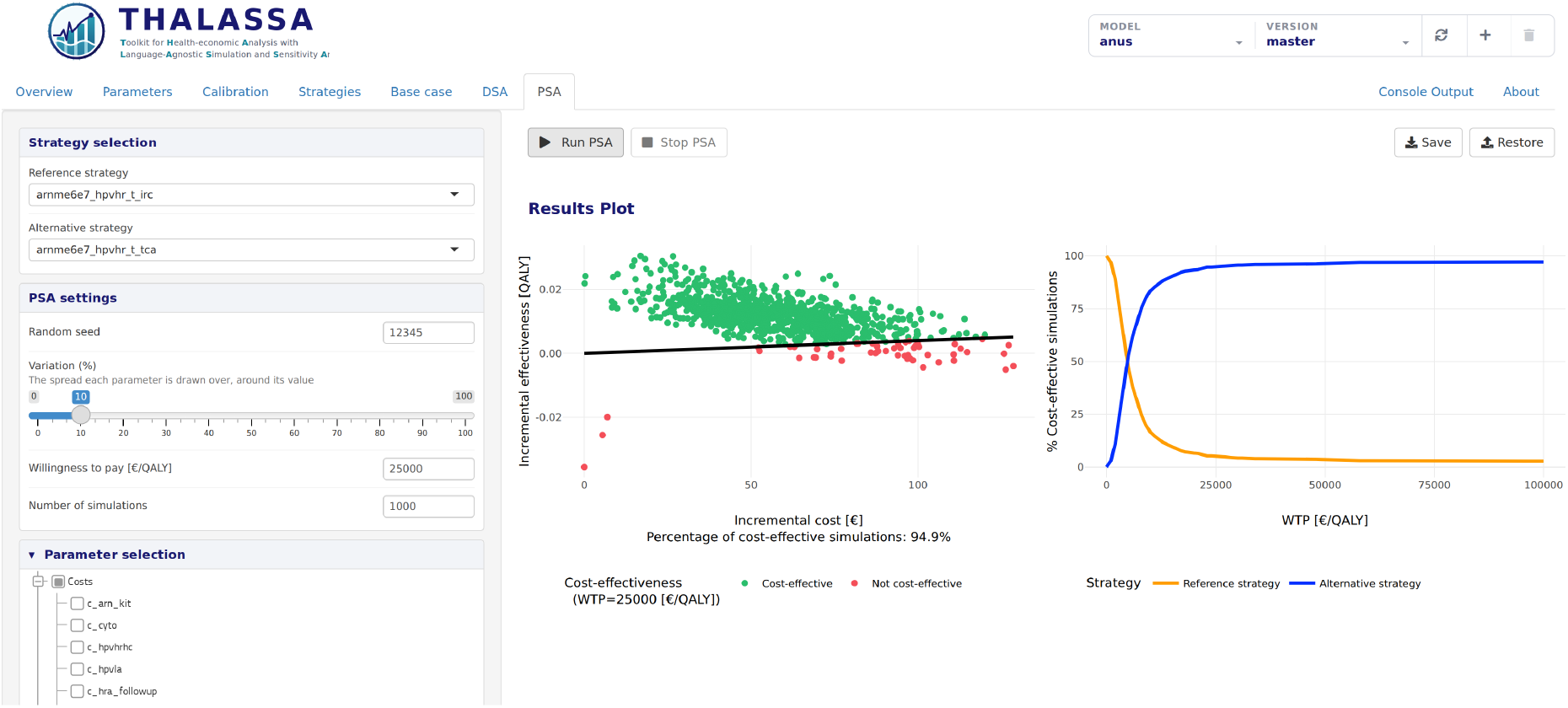
The PSA tab of case study 3, TCA against IRC over 1,000 draws.

### 3.4 Synthesis across the case studies

The three case studies share three results. First, no analysis code was written. Model inspection, parameter exploration, calibration, the base case and both sensitivity analyses were performed through the interface, and the exported workbooks are the reporting artifacts. Second, THALASSA itself was not modified. Connecting each model required implementing the same functions of Table 1, so the requirement placed on the model author was identical across the projects, although the second model carries more than six times as many parameter entries, four times as many states and a calibration of more than twice the dimension. Third, every reported result is traceable. Each figure and table above was produced by a tab bound to a pinned model version and saved to a workbook that the same tab reads back. Scenario questions raised during interpretation can be answered within the session by editing parameters and re-running the affected step, without returning to the model repository.

## 4 Discussion

The three case studies support a specific claim of practical importance. A single, unmodified analysis application can serve simulation models that share nothing except conformance to a small contract, and it can carry them through the complete CEA workflow. The two synthetic models were chosen to differ in what they demand of the user rather than only in subject, and the third shows the same workflow answering a real research question.

The benefit that users are likely to notice first is not calibration speed but the much lower cost of answering a follow-up question. Clinicians, health economists and decision makers who do not program can explore the model themselves. A question they raise can be answered within the same session, because parameter editing, re-calibration and all downstream analyses are interactive and changes propagate through them.

The reproducibility properties follow from a design decision rather than from analyst discipline. Because a session cannot begin without pinning a model version, and because every export carries the parameter values it used, the link between a reported estimate and its origin is maintained by default. Transparency and reporting guidance states this requirement explicitly [7, 8], whereas script-based workflows satisfy it only by convention. The second case study also shows why a choice of calibration methods matters. The default optimisation method is adequate for a simple model such as the first one, but not for a model of the complexity of the second. On that model it needed far more simulations than the other methods, and under constraints it wasted much of its budget on infeasible solutions. Because all methods run from the same tab, a user can move to a more advanced one without understanding its technical details.

Several existing tools address parts of this space. Dedicated decision-analytic software such as TreeAge [6] and Amua removes the scripting layer by hosting the model within the package itself. These are desktop applications built around their own model representations, so the model is confined to what the package supports and loses the flexibility of a general-purpose programming language. An existing model written in such a language must be rebuilt rather than connected. OCEAN [2] offers an accessible web interface for cervical cancer prevention analyses, but bundles one specific model, so the interface cannot be reused for a different disease. The `hesim R`package [5] provides a flexible and efficient modelling framework, but is a programming library rather than an interactive application, so it does not remove the scripting layer.

To our knowledge no available tool combines a language-agnostic contract for connecting an existing model, a complete interactive CEA workflow covering calibration through probabilistic sensitivity analysis, and version-bound result provenance. THALASSA is designed around that combination.

Future work will extend the uncertainty modules toward interpretability and explainability, applying attribution methods to probabilistic sensitivity analysis samples in order to expose higher-order parameter interactions that one-way analyses cannot reveal.

In conclusion, THALASSA provides the infrastructure surrounding cost-effectiveness analysis for any simulation model that implements a seven-function contract. It covers the complete workflow in the browser, from inspecting the model description and its parameters through calibration to probabilistic sensitivity analysis. A minimal model and a two-pathway cancer screening model were analysed end to end from the same installation, with no changes to the application and no analysis code. Calibration offers a range of methods, from traditional local search to state-of-the-art optimisation, that non-technical users can run and compare from one tab. Both models are public and a demo is available in the project documentation, so both case studies can be replicated. By binding every session to an explicit model version and exporting inputs alongside results, THALASSA makes the provenance of a reported estimate a property of the workflow rather than of the analyst s records. The result is a single reusable environment in which any conforming model can be calibrated, analysed and reported on without writing code.

## Data Availability

All data produced in the present study are available upon reasonable request to the authors

https://thalassa.iconcologia.net

## Funding

This work was supported by the Carlos III Health Institute (Spanish Government) through projects PI22/00219, as well as through CIBERESP CB06/02/007 (Consorcio de Investigación Biomédica en Red de España), co-financed by the European Union’s Horizon 2020 research and innovation program, a way to build Europe. Additional support was received from the Secretariat for Universities and Research of the Department of Business and Knowledge of the Government of Catalonia through grants to support the activities of research groups (SGR 2017-19 and 2021-23) (Grant numbers 2017SGR1718, 2017SGR1085, 2021SGR01029, and 2021SGR01354). Jesus Cerquides’ and Josep Lluís Arcos’ work was supported by grant PID2022-1 6787NB-I00 funded by MCIN/AEI/10.13039/501100011033.

## Conpeting interests

The authors declare that they have no competing interests.

## Acknowledgenents

We thank the CERCA Programme and Generalitat de Catalunya for institutional support.

## Notes

### Competing Interest Statement

The authors have declared no competing interest.

