## Appendix for "A Toolkit for Health-economic Analysis with Language-Agnostic Simulation and Sensitivity Analysis (THALASSA)"

This supplement gives the detail behind the main text. Appendix A documents the model integration contract and Appendix B the calibration methods. Appendices C and D give the calibration protocols of the two case studies, so that a reader can repeat them, and Appendix D also holds the calibration figures of case study 2. Appendix E describes the exported workbooks and how a result is reproduced from them, and Appendix F gives the full mapping to good-practice and reporting guidance. Both case-study models are public, at `https://github.com/david-gomez-guillen/cea-model-test` and `https://github.com/david-gomez-guillen/cea-model-test2`.

**C Case study 1 calibration results** **13**

**D Case study 2 calibration protocol and figures** **14**

**E Exported workbooks and reproduction** **15**

**F Good-practice and reporting guidance** **15**

### A The model integration contract

A model is connected by a file named `thalassa_interface.R` at the root of its repository. THALASSA sources this file after checking out the version the user selected, and from then on reaches the model only through the functions the file defines. The layout of the model directory, its implementation language and its structure are therefore irrelevant. Table 1 of the main text lists the functions. This appendix states what THALASSA calls each one with, what it expects back, and what happens when an optional one is absent.

#### A.1 Interface functions

`get.parameters()` Required. Takes no arguments and returns one descriptor per parameter. Each descriptor is a list holding the fields of Table A1, of which only the name and the base value are compulsory.

Table A1: The fields of a parameter descriptor. Only `name` and `base.value` are compulsory.

| Field | Meaning |
|---|---|
| `name` | The name of the parameter as used by `run.simulation()`, and the name a calibration scheme refers to it by. |
| `base.value` | The value the model runs on unless the user edits it or a calibration overrides it. |
| `display.name` | A readable label shown beside the name and searched together with it. Defaults to the name. |
| `stratum` | The stratum this descriptor holds the value for, naming one of the strata of `get.strata()`. Absent when unstratified. |
| `class` | The group name the parameter is shown under, such as costs, probabilities or utilities. Parameters declaring none are collected under a default heading. |
| `min.value`, `max.value` | Bounds enforced by the editor. A parameter declaring neither is left open. |
| `distribution` | The family the PSA draws from, one of beta, gamma, lognormal, normal and exponential, fitted to the base value and to a spread the user sets as a percentage of it. Defaults to normal. |
| `hidden` | Keeps the parameter out of the Parameters tab and out of the calibration, DSA and PSA parameter trees, while still passing it to every simulation at its base value. |

`get.strategies()` Required. Takes no arguments and returns the strategies, either as a vector of names, optionally with display labels, or as a named list that groups them under headings. The name is what `run.simulation()` receives. The label is what the selection tree and the results tables show. The grouped form drives the hierarchical tree that makes a large set of strategies navigable.

`get.strata()` Required of any stratified model. Takes no arguments and returns the names of the strata, for example age groups. These index stratified parameters and calibration targets. A model that declares none is unstratified throughout the simulation.

`run.simulation(strategies, pars)` Required. Called with the strategies to run and the parameter values to run them under. It returns a list containing a `summary` data frame with the columns `strategy`, `C` for cost and `E` for effectiveness, one row per strategy. Further elements may be returned alongside, for use by the calibration error. The values in `pars` are the base values with the user's edits and, when calibration is active, the calibrated values over them. A parameter arrives as a single value or as a list keyed by stratum.

`get.calibration.schemes()` Required for calibration. Takes no arguments and returns a named list of schemes, described in Appendix A.2. Without this function the Calibration tab is unavailable.

`get.overview()` Optional. Returns markdown describing the model, which the Overview tab shows. When absent, THALASSA generates an overview from the rest of the interface: the strategies, the strata, the parameters by class with their base values, and each calibration scheme.

`get.model.settings()` Optional. Returns model-wide settings, currently three. The cost and effectiveness units label the tables, plots and exported workbooks of the Base case, DSA and PSA tabs, and the willingness-to-pay threshold is read as a cost per effectiveness unit. `run.initial.guess` decides whether the Calibration tab simulates the initial guess as soon as it has a scheme to show, and a model too slow to be run on merely opening the tab sets it to false, which leaves that first simulation to the start of a calibration. Any setting left out falls back to the installation defaults.

#### A.2 Calibration schemes

A scheme is where the model author records the calibration decisions that he believes are appropriate for the model. Table A2 lists every field a scheme may declare. Five fields are needed for a scheme to run. Each of the others enables one capability of the Calibration tab and is ignored when absent.

Table A2: The fields of a calibration scheme. A scheme is one entry of the named list returned by `get.calibration.schemes()`.

| **Field** | **Status** | **Meaning** |
|---|---|---|
| `description` | Required | A sentence naming the scheme, shown in the scheme selector |
| `parameters` | Required | The parameters left free. Every other parameter is held at the value the Parameters tab supplies |
| `strata` | Required | The strata the free parameters are indexed by, which fixes the layout of the calibration vector |
| `target` | Required | A named list of target series, each mapping stratum names to observed values |
| `initial_guess` | Optional | The point the search starts from. Without it the search starts from the parameter values currently in force |
| `error_function` | Required | The discrepancy measure. It receives the parameter set and the target and returns the error together with the model quantities compared against the target. THALASSA does not interpret the error scale |
| `constraints` | For constrained | A function returning one violation per requirement, negative when satisfied. Declaring it makes the constraint-aware methods of Appendix B available and makes the error plot shade infeasible simulations |
| `constraints_description` | Optional | The same requirements as a sentence, shown beside the scheme and passed to the agent-driven calibrator |
| `latent_space_training_set` | Optional | A generator of plausible parameter sets, on which the latent spaces of Appendix B are trained |
| `other.plots` | Optional | Model-specific diagnostic plots drawn beside the standard ones |

#### A.3 Example implementation

The listing below is the interface file of the tutorial model of case study 1, taken from `cea-model-test` and reproduced in full, except that the commented-out test block at its end is left out. A comment on `run.simulation()`, a `get.model.settings()` and a constrained calibration scheme with the two functions it needs are added here, and the model itself declares none of them.

```
library(ggplot2)

source('model.R')

get.overview <- function() {
  # Markdown shown in the Overview tab.
  return(paste(readLines('overview.md'), collapse='\n'))
}

get.model.settings <- function() {
  # The units that label every result table, plot and exported workbook, and
  # whether the Calibration tab simulates the initial guess as soon as it is
  # opened, which a model too slow for that would leave to the start of a
  # calibration. Settings left out fall back to the installation defaults.
  return(list(
    cost.unit='€',
    effectiveness.unit='QALY',
    run.initial.guess=TRUE
  ))
}

get.strategies <- function() {
  # Hardcoded strategies for the model. In a real application, these could be loaded
      from a file or database.
  return(list(
    list(name='no_intervention', display.name='No intervention'),
    list(name='screening', display.name='Screening'),
    list(name='treatment', display.name='Treatment')
  ))
}

get.parameters <- function() {
  # Hardcoded parameters for the model. In a real application, these could be loaded
      from a file or database.
  return(list(
    list(
      name='p.healthy.cancer',
      display.name='Annual probability of developing cancer while healthy',
      base.value=0.13,
      class='General'
    ),
    list(
      name='p.healthy.death',
      display.name='Annual probability of death while healthy',
```

```
    base.value=0.00001,
    class='General'
  ),
  list(
    name='p.cancer.death',
    display.name='Annual probability of death while having cancer',
    base.value=0.0001,
    class='General'
  ),
  list(
    name='p.cancer.recovery',
    display.name='Annual probability of cancer resolving on its own (back to healthy)
        ',
    base.value=0.3,
    class='General'
  ),
  list(
    name='p.screening.effective',
    display.name='Proportion of cancer cases that are prevented by screening',
    base.value=0.05,
    class='Screening'
  ),
  list(
    name='p.treatment.effective',
    display.name='Annual probability of the regular treatment curing cancer (back to
        healthy)',
    base.value=0.03,
    class='Treatment'
  ),
  list(
    name='cost.screening',
    display.name='Annual cost per healthy person under screening',
    base.value=15000,
    class='Screening'
  ),
  list(
    name='cost.cancer.treatment',
    display.name='Annual cost per person with cancer under regular treatment',
    base.value=200000,
    class='Treatment'
  ),
  list(
    name='utility.cancer',
    display.name='Utility of a year spent with cancer',
    base.value=0.6,
    class='General'
  ),
```

```
    list(
      name='discount',
      display.name='Discount rate',
      base.value=0.03,
      class='General'
    )
  ))
}

get.strata <- function() {
  # Hardcoded strata for the model. In a real application, these could be loaded from
      a file or database.
  return(c('30-34', '35-39', '40-44', '45-49', '50-54', '55-59', '60-64', '65-69', '
      70-74'))
}

run.simulation <- function(strategies, pars) {
  # The pars vector should be transformed to the format expected by the simulate
      function.
  # This is a simple mapping based on the parameter names.
  # This mapping is specific to how this model is written. Another model would
  # translate the same pars into whatever form it is called in, be it a function
  # signature, a configuration file or an executable in another language.
  results <- simulate(strategies,
                     p.healthy.cancer=pars[['p.healthy.cancer']],
                     p.healthy.death=pars[['p.healthy.death']],
                     p.cancer.death=pars[['p.cancer.death']],
                     p.cancer.recovery=pars[['p.cancer.recovery']],
                     p.screening.effective=pars[['p.screening.effective']],
                     p.treatment.effective=pars[['p.treatment.effective']],
                     cost.screening=pars[['cost.screening']],
                     cost.cancer.treatment=pars[['cost.cancer.treatment']],
                     utility.cancer=pars[['utility.cancer']],
                     discount=pars[['discount']])
  return(results)
}

get.calibration.schemes <- function() {
  return(list(
    standard=list(
      description='Example calibration',
      parameters='p.healthy.cancer',
      target=list(
        # Each target is a named list assigning a value to a specific stratum.
        # Strata not listed here are not calibrated against (e.g. burn-in strata).
        `Cancer incidence`=list(
          `30-34`=.01,
```

```
          `35-39`=.05,
          `40-44`=.08,
          `45-49`=.1,
          `50-54`=.11,
          `55-59`=.12,
          `60-64`=.13,
          `65-69`=.135,
          `70-74`=.14
        )
      ),
      strata=get.strata(),
      initial_guess=rep(.13, 9),
      error_function=calibration.error,
      latent_space_training_set=generate.training.dataset,
      other.plots=NULL
    ),
    # A constrained scheme is an ordinary scheme with the two fields that state
    # its constraints.
    constrained=list(
      description='Example calibration, incidence non-decreasing with age',
      parameters='p.healthy.cancer',
      target=list(
        `Cancer incidence`=list(`30-34`=.01, `35-39`=.05, `40-44`=.08,
                                `45-49`=.1, `50-54`=.11, `55-59`=.12,
                                `60-64`=.13, `65-69`=.135, `70-74`=.14)
      ),
      strata=get.strata(),
      initial_guess=rep(.13, 9),
      error_function=calibration.error,
      constraints=calibration.constraints,
      constraints_description=calibration.constraints.llm,
      latent_space_training_set=generate.training.dataset,
      other.plots=NULL
    )))
}

# Constraints follow the c(x) <= 0 convention, one value
# per requirement, so a value is negative when the requirement is met. They are
# evaluated on the raw calibration vector and the requirement in this example is that
    the
# annual probability of developing cancer does not fall with age.
# `partial` says the vector is only filled in as far as a stepwise calibration
# has got, so the first step, which holds a single value and no pair to compare,
# is given a constant that is satisfied by construction.
calibration.constraints <- function(x, partial=FALSE) {
  x <- as.numeric(x)
  if (length(x) < 2) return(-1)
```

```
  return(head(x, -1) - tail(x, -1))
}

# The same requirement in words, which the agent-driven
# calibrator calls with no arguments and appends to its instructions.
calibration.constraints.llm <- function() {
  paste0('The vector holds p.healthy.cancer as one value per age group, in the ',
         'order ', paste(get.strata(), collapse=', '), '.\n',
         'The values are non-decreasing: no value is smaller than the one ',
         'before it.')
}

calibration.error <- function(pars, target) {
  calibration.strategy <- 'no_intervention'
  # The target is a named list with one entry per stratum, so it is flattened into
  # a named numeric vector to match the simulated values by stratum name.
  target.inc <- unlist(target$`Cancer incidence`)
  result <- tryCatch({
    results <- run.simulation(calibration.strategy, pars)
    cancer.incidence <- results$incidence[[calibration.strategy]]
    names(cancer.incidence) <- get.strata()
    # Only the strata present in the target contribute to the error.
    error <- sum((cancer.incidence[names(target.inc)]-target.inc)^2)
    result <- list(
      error=error,
      output=list(cancer.incidence=cancer.incidence)
    )
    result
  }, error=function(e) {
    error <- Inf
    cancer.incidence <- rep(NA, length(get.strata()))
    names(cancer.incidence) <- get.strata()
    result <- list(
      error=error,
      output=list(cancer.incidence=cancer.incidence)
    )
    result
  })
  return(result)
}

generate.training.dataset <- function(initial_guess, n, ...) {
  # The training set of the constrained scheme. It returns n parameter sets the model
      author
  # considers plausible, one per row, on which the latent space of the
  # latent-space methods is trained, so that the search moves over sets of this
  # kind rather than over the raw parameter space. Here they are drawn by
```

```
  # scaling the initial guess by a random factor, whose width is the search
  # scope the user sets in the Calibration tab, which arrives through `...` as
  # `variation`.
  f.pars <- list(...)
  variation <- f.pars$variation

  n_params <- length(initial_guess)

  dataset <- matrix(NA, nrow=n, ncol=n_params)

  for(i in 1:n) {
    factors <- runif(n_params, min=1-variation, max=1+variation)
    dataset[i,] <- pmin(1, initial_guess * factors)
  }

  dataset <- dataset[sample(nrow(dataset)),]

  return(dataset)
}
```

### B Calibration methods

Most users meet calibration as a configured step: they select a scheme, set a method, a budget and run it. This appendix describes the options underneath, for models difficult enough to need them. Their development and evaluation are reported separately [5; 6].

#### B.1 Why a generic optimiser is not enough

Each evaluation of the calibration error is a full model run, which takes seconds or minutes for a production-scale model. Methods that need tens of thousands of evaluations are then impractical. Bayesian optimisation addresses this with a probabilistic surrogate of the error, choosing each next evaluation where it expects the most progress [4]. Both case-study models are fast to compute, so both repositories add an artificial delay to each simulation, to reproduce the conditions the calibration module is designed for.

A low error is also not sufficient. A parameter set may reproduce the observed targets while assuming a solution that is not clinically credible. The unconstrained runs of case study 2 return exactly that (Figure D1). Calibration must therefore also be steered towards plausible regions.

#### B.2 Optimisation algorithms

Table B1 lists the available algorithms. They are options of one selector in the Calibration tab, so switching between them needs no change to the model or the scheme. Standard Bayesian optimisation is the default.

Table B1: Optimisation algorithms available in the Calibration tab.

| **Algorithm** | **Intended use** |
|---|---|
| Standard BO | Gaussian-process surrogate with a standard acquisition function. The default, adequate for low-dimensional problems such as the calibration of case study 1. |
| TuRBO | Trust-region BO [10], for problems where a global surrogate fits poorly. In the 21-dimensional calibration of case study 2 it reached an error about 30 times lower than standard BO on the same budget. |
| BAxUS | Adaptive nested subspaces [11], for higher-dimensional problems where only a few directions matter. |
| SAASBO | Sparse axis-aligned subspace priors [12], for high-dimensional problems where few parameters have a strong effect. |
| Nelder–Mead | Derivative-free local search, kept as a baseline and for refinement from a good starting point. |
| Simulated annealing | Stochastic local search, a second non-Bayesian baseline. |
| SCBO | Constrained trust-region BO [13], which takes the constraints into account when choosing each evaluation. Offered only for a scheme that declares constraints. |
| Agent-driven (LLM) | A large language model proposes each next parameter vector from the history of simulations [6], and reads constraints stated in plain language. |

#### B.3 Latent-space embeddings

Instead of searching the full parameter space, the module can learn a low-dimensional representation from plausible parameter sets supplied by the model, and search within it [7]. This reduces the search dimension and keeps proposals close to the plausible sets the representation was trained on. The available mappings are principal component analysis, autoencoders, variational autoencoders (VAE) [8] and factorised bijective flows, or none. The training set comes from the generator the scheme declares, which is where the model author states what counts as plausible. In case study 2, Nelder–Mead in a VAE trained on feasible sets reached an error about 15 times lower than Nelder–Mead penalising infeasible proposals, with two thirds of the simulations and no infeasible proposal (Table 5 of the main text).

##### B.4 Constraints

Constraints belong to the scheme, not to the method. A Bayesian method rejects an infeasible candidate and draws another, or, with *Use constraint modeling* ticked, models the probability that a candidate is feasible and weights its choice by it [9]. Nelder–Mead and simulated annealing instead add a penalty to the error of infeasible proposals. SCBO and the agent handle constraints themselves. A stepwise option, shown as a checkbox beside the method selector, calibrates groups of parameters in sequence rather than jointly [5], and each step is held to the constraints that the parameters calibrated so far already decide. The error plot shades simulations that violated the constraints, so the user sees whether a method is spending its budget inside the feasible region.

##### B.5 Agent-driven calibration

The agent-driven method replaces the surrogate with a large language model that proposes each next parameter vector [6]. The agent sees the history of simulations, the parameter bounds and, if the user ticks the option, a summary of the model's source code. When the scheme declares a `constraints_description`, that sentence is added to the agent's instructions, and every proposal is checked against the numeric constraints before it is simulated. The agent needs far fewer simulations than any other method, 25 in case study 2 against 110 to 1,320. Its rationale for each proposal is streamed to the console tab and kept in a run log.

#### C Case study 1 calibration results

`model_test`, pinned at branch `main`, declares three strategies, ten unstratified parameters in three classes, nine five-year age strata from 30 to 74, and one calibration scheme. All values below are those shown in the figures of the main text.

| | | # Simulations | | | |
|---|---|---|---|---|---|
| **Method** | **Iteration budget** | **Initial** | **Search** | **Elapsed** | **Best error** |
| Nelder–Mead | 100 | 10 | 140 | 1:05 | $2.12\times10^{-4}$ |
| Bayesian optimisation | 100 | 10 | 100 | 2:16 | $1.17\times10^{-6}$ |

Table C1: The two calibration runs of case study 1, as reported by the Calibration tab. The iteration budget is the number the user sets. The simulations are the model runs spent, split into those of the initial design, drawn before the search begins, and those of the search itself. For Nelder–Mead the initial design is the starting simplex, one point more than the nine free parameters, as in case study 2. Nelder–Mead spends a variable number of runs per iteration, so its total, 150, is read off the evaluation axis of Figure 6a of the main text.

Figure C1 shows the calibration plots at the end of each run. In both panels, the left plot compares the simulated incidence curve, dashed, with the target, solid, and the right plot is the error trace, whose last evaluation is marked.

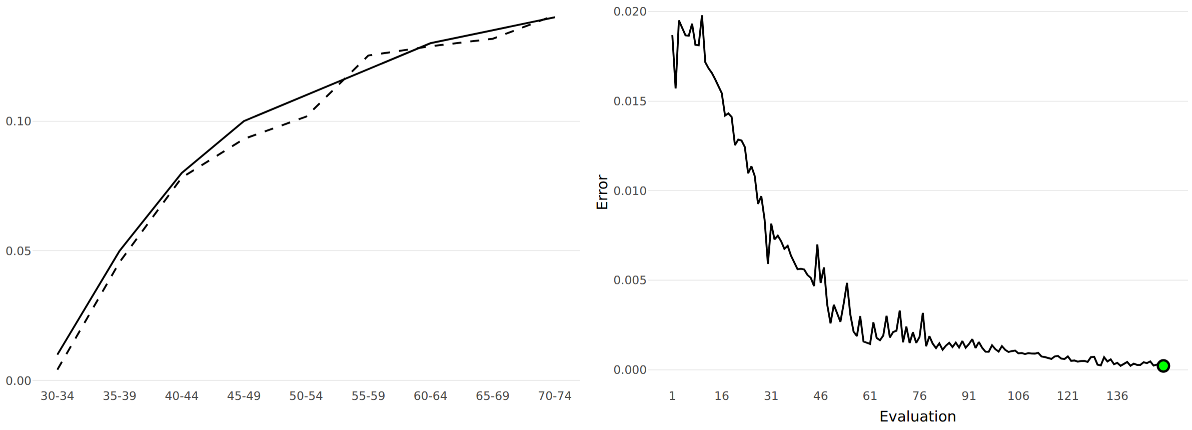

(a) Nelder–Mead

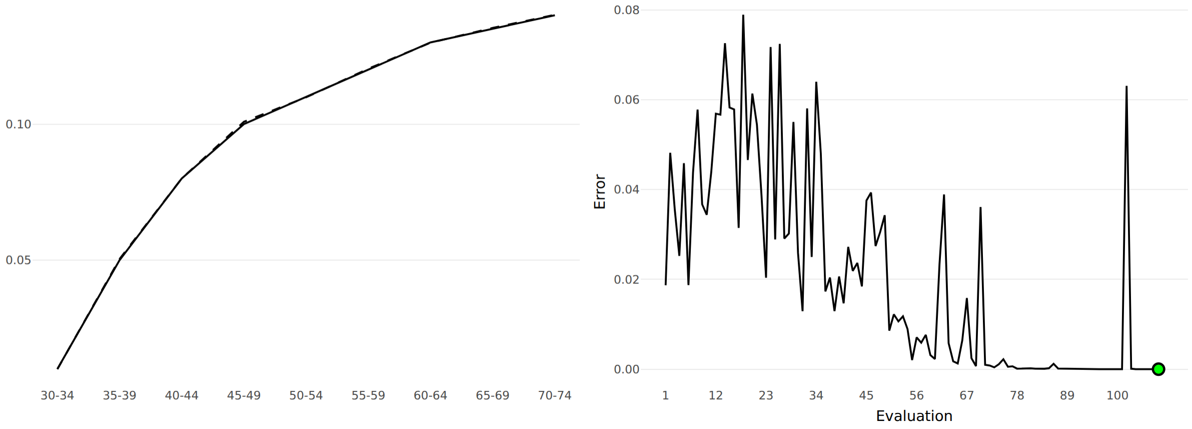

(b) Bayesian optimisation

Figure C1: The calibration plots of case study 1 at the end of each of the two runs of Table C1, under the *Example calibration* scheme. Nelder–Mead leaves the simulated curve below the target up to 54 and above it at 55–59, where the Bayesian run makes the two curves visually indistinguishable.

### D Case study 2 calibration protocol and figures

`model_test2`, pinned at branch `master`, declares four strategies grouped under three screening modalities, 64 parameter entries over 46 distinct names in nine classes, seven ten-year age strata from 20 to 89, and two calibration schemes.

Figures D1 and D2 show the Calibration tab at the end of each run. In every panel, the top left plot compares the three simulated series with their targets, the top right plot is the error curve, and the bottom plot shows the calibrated onset probabilities of the two lesion types by age group.

### E Exported workbooks and reproduction

Each analysis tab saves its run as a timestamped workbook, with the contents listed in Table E1. Every workbook carries a *Base parameters* sheet with the complete parameter set the run used, one row per parameter and stratum, marking which values came from a calibration. The values are those in force when the run started, not those on screen when the file is downloaded. Every workbook also carries a *Settings* sheet with the model and version, the time the file was written and the settings the run was made with. The Parameters tab writes and reads a workbook of its own, which is how a parameter set is carried between sessions.

Table E1: Contents of the exported workbooks, in addition to the parameter and settings sheets that every workbook carries.

| **Tab** | **Exported content** |
|---|---|
| Calibration | Calibrated parameter values, targets, error trace over the evaluations, and the scheme and method settings. |
| Base case | Cost and effect per strategy, incremental analysis, the ICER, which is absent for dominated strategies, and the NHB at the threshold in force. |
| DSA | Low and high results per parameter, the variation applied, and the ranking behind the tornado chart. |
| PSA | Sampled parameter values, results per sample, and the acceptability curve by willingness-to-pay threshold. |

### F Good-practice and reporting guidance

Table F1 extends Table 2 of the main text. It is not a claim that using THALASSA satisfies these guidelines, since most items concern modelling decisions that THALASSA cannot make. It shows which items the workflow supports by construction and which remain the analyst's responsibility.

Table F1: Mapping to the ISPOR-SMDM Modeling Good Research Practices [1; 2] and CHEERS 2022 [3].

| **Requirement** | **Status** | **Mechanism or gap** |
|---|---|---|
| Model version identifiable for any reported result | Enforced | A session cannot start without selecting a branch or tag |

| Requirement | Status | Mechanism or gap |
|---|---|---|
| Parameter values reported with results | Produced | Parameter sheet in every exported workbook, marking calibrated values |
| Parameter sources and justification | Analyst | THALASSA records values, not their sources. The model's overview is where the author can state them |
| Model structure and assumptions described | Supported | Overview tab, from the model's own description or generated from the interface |
| Calibration targets stated | Produced | Declared per stratum in the scheme and exported with the results |
| Calibration method and criteria stated | Produced | Scheme, method, budget and error trace exported |
| Calibration identifiability assessed | Analyst | Not assessed by THALASSA |
| Structural assumptions stated | Analyst | Kept in the model repository |
| Strategies compared and dominance reported | Produced | Efficiency frontier, with dominated strategies reported without an ICER |
| Decision threshold stated with results | Supported | Willingness-to-pay input on the Base case, DSA and PSA tabs, with the NHB reported at it |
| Costs and effects reported in stated units | Produced | Units declared by the model label every table, plot and workbook |
| Discounting applied and reported | Supported | Exposed as a model parameter where the model declares it |
| Deterministic sensitivity analysis reported | Supported | Available for any conforming model |
| Probabilistic sensitivity analysis reported | Supported | Uses the distributions declared by the model |
| Distributional assumptions for PSA | Analyst | Declared per parameter in the interface file |
| Model validation, internal and external | Analyst | THALASSA does not validate the model it loads |
| Analysis reproducible by a third party | Supported | Version plus exported workbook are sufficient, see Appendix E |
| Model internals inspectable during analysis | Supported | Live console stream and calibration diagnostics |

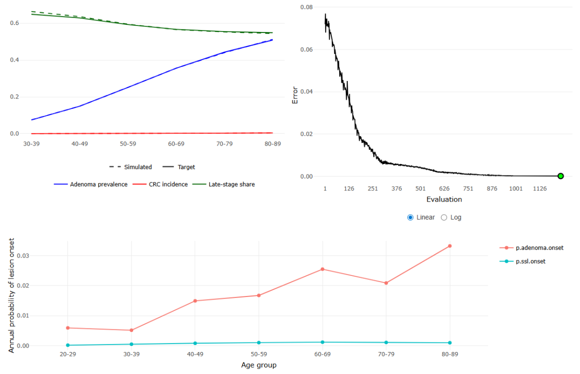

(a) Nelder–Mead

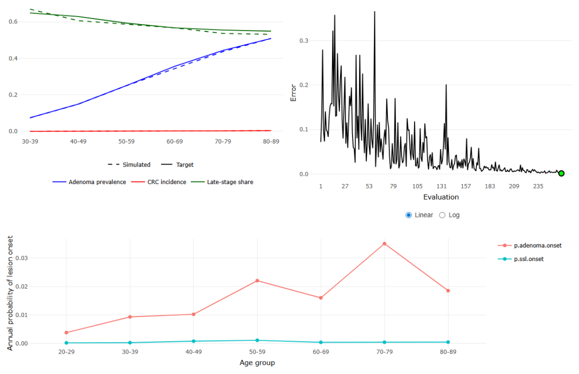

(b) Bayesian optimisation

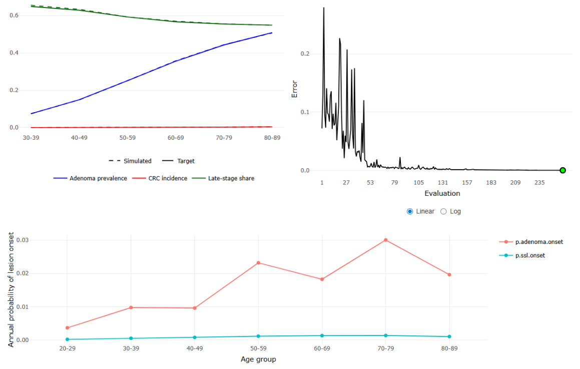

(c) TuRBO

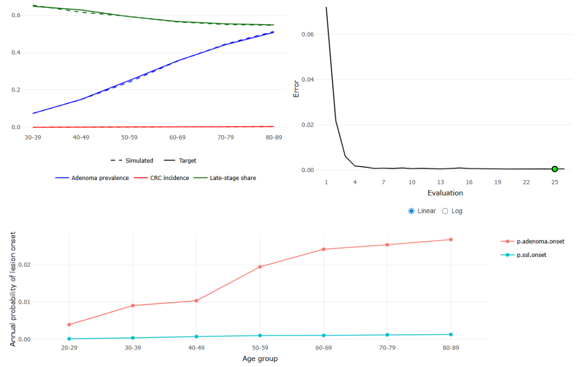

(d) Agent-driven calibration

Figure D1: The Calibration tab of case study 2 at the end of each of the four unconstrained runs of Table 4 of the main text, under the *Lesion onset and progression* scheme.

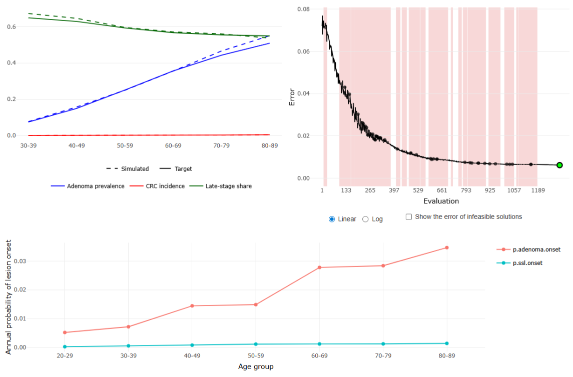

(a) Nelder–Mead with infeasible proposals penalised

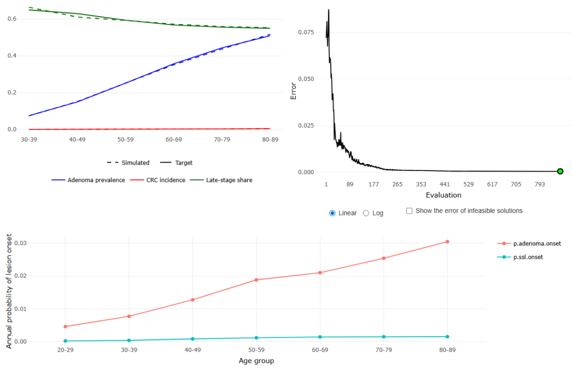

(b) Nelder–Mead in a VAE latent space

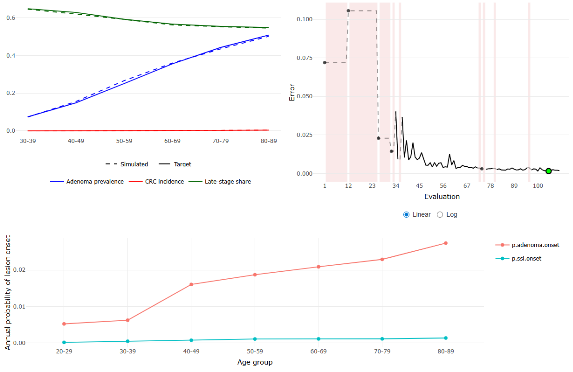

(c) SCBO

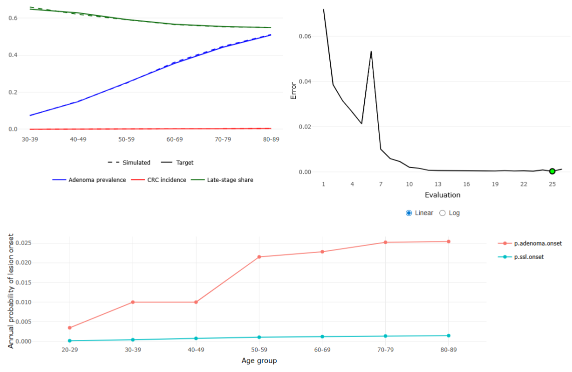

(d) Agent-driven calibration

Figure D2: The Calibration tab of case study 2 at the end of each of the four constrained runs of Table 5 of the main text, under the *Constrained onset and progression* scheme. The error plot of each panel shades the infeasible evaluations, and the onset curves are non-decreasing in age, as the scheme requires.
